# Hormones and Infant Caregiving in Postpartum Opioid Use Disorder Recovery: Compliance and Satisfaction with a Prospective Cohort Study Protocol Designed to Identify Novel Support Targets

**DOI:** 10.1101/2024.12.14.24319034

**Authors:** Alicia M. Allen, Linnea B. Linde-Krieger, Jendar Deschenes, Stephanie Mallahan, Alexandra Harris, Mariana Felix, Arushi Chalke, Alma Anderson, Priyanka Sharma, Katherine M. King, Maddy T. Grant, Lela Rankin, Stacey Tecot

**Author notes:** <u>Corresponding Author:</u> Alicia Allen, Department of Family and Community Medicine, College of Medicine – Tucson, University of Arizona, Tucson, AZ, USA, Mailing address: 3950 South Country Club Road, Suite 330, Tucson, AZ 85714, USA.

## Abstract

**Background:** Although treatment for opioid use disorder (OUD) often yields high adherence during pregnancy, the risk of returning to opioid misuse during postpartum is high. There are currently no relapse prevention programs tailored to this unique time period.

**Objectives:** Using a prospective cohort study, we seek to identify hormones and/or infant caregiving approaches as predictors of postpartum opioid misuse. As a first step in dissemination of results, this report contains a detailed account of the protocol, as well as recruitment, retention, compliance, and participant satisfaction.

**Population:** Participants were individuals with OUD (OUD+) and those without (OUD−) who were followed from late pregnancy (≥gestational week 36) to postpartum month five.

**Methods:** From childbirth to postpartum week 12, participants completed daily surveys (capturing use, craving, interactions with infant) and weekly face-to-face visits (including collection of biological samples for hormone assays). Follow-up visits using the same procedures occurred at postpartum month four and five.

**Preliminary Results:** A total of 50 OUD+ and 21 OUD− participants enrolled, 93% notified the study staff of childbirth, 87% completed at least one postpartum clinic visit, and 73% completed through postpartum month five. Compliance with procedures ranged from 66% for saliva samples and daily surveys to 78% for weekly surveys, generally with lower compliance among OUD+ and at later study time points. Participants, regardless of group and time point, reported high study satisfaction (e.g., on a scale where 0 is “not at all” and 3 is “extremely”, on average participants reported 2.9±0.4 for their willingness to complete this study again).

**Conclusions:** This prospective cohort study was well tolerated despite the challenging postpartum period. Data collected will provide ample opportunities to identify novel risk/protective factors to inform the development of new relapse prevention intervention programs specific to the needs of those with OUD during early postpartum.

**Synopsis:** Using a novel protocol with daily surveys plus weekly face-to-face visits with biological sample collection from late pregnancy through 12 weeks postpartum, we report on the compliance with data collection methodologies over time between those with and without opioid use disorder. We observed fairly high levels of retention (e.g., 75% completed through postpartum week 12) and compliance (e.g., 69% of weekly dried blood spots were collected) with very high satisfaction (e.g., on a scale where 0 is “not at all” and 3 is “extremely”, on average, participants reported 2.9±0.4 for their willingness to participate again). These observations suggest that both those with and without opioid use disorder are willing and able to participate in an intensive, multi-method, prospective study during the early postpartum period.

## Background

Perinatal opioid use disorder (OUD) has increased by 600% from 1.5 per 1,000 hospitalized deliveries in 1999 to 8.2 per 1,000 hospitalized deliveries in 2017.^1,2^ Opioid overdose is now a leading cause of maternal mortality,^3^ with additional significant adverse impacts on the birthing person, infant, family, and beyond.^4,5^ For example, those with OUD are more likely to have their infants removed by child protective services, heightening risk for postpartum depression and anxiety, disrupted parent-child attachment, and impaired child socioemotional development.^6–8^ Preventing postpartum return to opioid misuse would not only prevent fatal maternal overdoses, but also mitigate numerous adverse consequences.

Medication for OUD (MOUD) is the gold standard treatment of perinatal OUD, improving both maternal and infant outcomes.^9–11^ However, the risk for return to opioid misuse and overdose increases during postpartum.^12,13^ To address this period of high-risk, the American College of Obstetricians and Gynecologists and the American Society of Addiction Medicine, recommends that “*Access to…relapse prevention programs should be made available.*”^11^ However, there are currently no evidence-based relapse prevention programs tailored to the unique postpartum circumstances.^14^ Additionally, factors that contribute to the increased risk of return to use during postpartum are not well understood.^3^ Ultimately, developing effective relapse prevention programs tailored to postpartum-specific needs will allow parents, infants, and families enhanced opportunities to achieve positive long-term outcomes.^3^

Biological females are more vulnerable to substance use disorder development, maintenance, and relapse.^15,16^ Sex differences, potentially driven by ovarian hormones, have important implications for clinical treatment.^15–17^ For instance, in ovariectomized female rats offered intermittent access to fentanyl, those who received exogenous estradiol treatment exhibited increases in self-administration, enhanced sensitivity, and increased motivation versus the placebo treatment.^18^ Research by our group and others has also indicated that exogenous progesterone treatment is associated with reductions in combustible cigarette smoking in pregnant and non-pregnant females.^19–21^ Hormonal influences on substance misuse behaviors may be amplified during the perinatal period, as hormones change by up to 300-fold during pregnancy followed by a dramatic decline early postpartum and additional variation with lactation.^22^ Interestingly, some of the natural hormonal patterns that occur during the perinatal period directly mirrors the risk of substance misuse. For instance, both estrogen (which facilitates drug-taking behaviors) and risk of substance misuse are low during lactation whereas without lactation both estrogen and risk of substance misuse increase.^22,23^ Further, specific infant caregiving activities can have substantial effect on hormones (e.g., oxytocin increases during skin-to-skin contact) which may directly or indirectly impact postpartum substance relapse risk.^24–27^ It is currently unknown if specific infant caregiving activities may provide a unique opportunity to protect against postpartum return to opioid misuse.

We conducted a prospective cohort study with those who do and do not have perinatal OUD to examine the utility of hormones and/or infant caregiving activities to protect against return to opioid misuse during the high-risk postpartum period. As a first step in dissemination, this paper aims to: (a) detail our protocols and methodologies, and (b) report on recruitment, retention, compliance, and participant satisfaction. Ultimately, this research will enhance the understanding of perinatal OUD to inform to the development of novel intervention and treatment options to support postpartum recovery and wellbeing.

## Methods

### Study Design and Study Sample

This study was approved by the University of Arizona’s Institutional Review Board. In this prospective cohort study, we sought to enroll 50 pregnant people with OUD (OUD+) and 25 pregnant people without OUD (OUD−) to ensure a final sample size of 60 (40 OUD+, 20 OUD−). Participants met the following inclusion criteria: (a) 18-40 years old, (b) uncomplicated single-gestation pregnancy at gestational week 30 or beyond, (c) a self-reported expectation of residing with the infant after birth, (d) English fluency, and (e) willing/able to comply with procedures. Additionally, OUD+ had to: (f) report use of opioids during pregnancy, (g) report current OUD treatment with plans to continue treatment after childbirth, and (h) allow us access to the OUD treatment records. All participants were excluded for preterm birth (<36 gestational weeks) or planned long-distance moves and/or extended travel through six months postpartum. Additional OUD− exclusion criteria included the use of opioids during pregnancy or a history of substance use disorder(s). Finally, we utilized frequency matching to increase exchangeability between study groups based on health insurance status (i.e., no health insurance or public insurance versus private insurance) and age (i.e., ± 5 years).

We recruited participants in the metro areas of Tucson and Phoenix, Arizona, from August 2021 to March 2024 via flyers/tabling at local healthcare provider clinics, community resources, community events, and social media. Individuals expressed their interest via a survey on REDCap,^28^ this was followed by a telephone-based eligibility interview with staff. If eligibility criteria were met, potential participants were invited to a face-to-face baseline visit to commence participation.

### Data Collection Procedures

Participants completed 15 visits either in-person (at our clinic or the participant’s home) or remotely via HIPAA-compliant Zoom for Health, with all data entered on REDCap.^28^ Visits were scheduled to begin between 7am and 11am given the known diurnal patterns in hormones and other variables of interest.^29,30^ For those who attended remotely, study supplies were mailed in advance. All visits included collection of biological samples with all participants provided 1.8 mL of saliva via passive drool. Remote participants self-collected 12 drops of blood for dried blood spots using our previously developed protocol.^31^ In-person participants provided 8 mL of venous blood, which was then processed to yield 12 drops of dried blood spots plus plasma samples. For remote participants, dried blood spots were mailed to staff weekly and frozen saliva samples were picked up in batches by staff. All samples were subsequently stored at ≤ −20°C.

#### Baseline

At gestational week 36 or beyond, participants completed a baseline visit. After providing informed consent, participants completed an interview with staff to obtain their medical history followed by completion of surveys, and biological sample collection. Next, participants completed a semi-structured audio-recorded interview with staff on lifetime trauma and resilience (Supplementary Document 1). At the end of the visit, participants were trained on how to complete daily surveys and at-home saliva samples followed by receipt of compensation.

#### Daily Surveys

Beginning the day after baseline through postpartum week 12, participants completed daily surveys on their own or study-supplied smartphones. This was replicated the week preceding follow-up visits. Participants received a daily text message or email at 8pm containing a link to surveys. Surveys remained open until 11:59pm, with reminders every 30 minutes until completed. Surveys took approximately 10-15 minutes to complete and documented use of and craving for substances, interactions and connectedness with infant and others, and other validated measures to capture return to use risk factors.

#### Fourth Trimester (Postpartum Weeks 1-12)

Participants notified staff of childbirth via the daily surveys then commenced weekly visits. At each visit, participants completed surveys and provided in-visit biological samples. Additional at-home saliva samples collected at 8pm the day before and 30 minutes after waking on the day of the visit. Participants were also interviewed to capture substance use, changes in health/medication, and time spent with infant. At postpartum week 1, participants completed a semi-structured audio-recorded interview with staff to capture birth and initial breastfeeding experiences (Supplementary Document 2). Approximately one year into study recruitment, we added a study satisfaction survey to postpartum weeks 1 and 12. Upon completion of each visit, the next visit was confirmed, and participants were compensated.

#### Follow-Up Visits (Postpartum Months 4 and 5)

Procedures were identical to the fourth trimester data collection. In addition, at postpartum month 5, participants completed a semi-structured audio-recorded interview with staff to capture their breastfeeding experiences and a final satisfaction survey.

### Study Measures

Overall, participants completed a series of instruments on a daily, weekly, and monthly basis, as well as at follow-up (Supplementary Table 1, Supplementary Document 3).

#### Background Variables

During the enrollment interview and at the baseline visit, standardized questionnaires and interviews were used to capture sociodemographics (e.g., age, race/ethnicity), medical history (e.g., parity, diagnosis of hormone-influencing conditions), history of substance use (e.g., lifetime use of 16 different substances, drug of choice, OUD treatment experiences). The baseline interview incorporated Stressful Life Events,^32^ Adverse Childhood Experiences,^33^ Early Trauma Inventory,^34,35^ and Resilience Scale.^36^

#### Maternal Factors

These variables included: (a) known/suspected risk factors for postpartum substance misuse, (b) known/suspected association with hormones of interest, and/or (c) infant caregiving and/or parental experiences. In brief, participants reported on lactation/breastfeeding (daily first four weeks, weekly thereafter), pain (daily first four weeks), and vaginal bleeding (daily after postpartum week four), with additional assessments on a weekly or monthly basis.

#### Caregiving Factors

Variables included those that may contribute to risk of postpartum return to misuse and/or influence hormone levels. Daily participants reported on their perceptions of parenting challenge, parenting reward, and connections with infant and with others using a 100-point visual analog scale ranging from “not at all” to “extremely.” Relative time spent with infant each day was reported using an investigator-created instrument with 100-point visual analog scale ranging from “a lot less than usual” to “a lot more than usual” to report on the amount of time spent: (a) in skin-to-skin contact with infant, (b) in other physical contact with infant, (c) caring for infant but not in physical contact, (d) with someone else while they cared for infant, and (e) time away from infant. Weekly staff interviewed participants to capture the average absolute time spent on each of these activities during the preceding week ensuring that total time reported equated to 24 hours per day. Additional validated items were completed regularly throughout follow-up.

#### Hormones

We measured a total of 19 hormones at each time point. Cortisol and oxytocin assays were completed by the Laboratory for the Evolutionary Endocrinology of Primates (LEEP) at the University of Arizona. Saliva samples were stored at −80°C, thawed to room temperature before analysis, and cleaned by centrifuging for 30 minutes at 3,000 rpm with the supernatant retained. Saliva was aliquoted into microcentrifuge tubes at volumes of 500 ul for oxytocin and 150 ul for cortisol then frozen at −80°C until lyophilization. Samples were lyophilized at −80°C and 0.3 mbar until completely dry, then resuspended in 250 ul and 150 ul of assay buffer (Arbor Assays, Ann Arbor, MI, US), respectively. This resulted in 2x concentration for oxytocin and 1x concentration for cortisol. Samples were assayed according to protocols included with DetectX ELISA oxytocin (K048-H5; cross reactivity with isotocin 94.3%, mesotocin 88.4%, other analytes <0.15%; sensitivity 17.0 pg/mL; limit of detection 22.9 pg/mL) and DetectX ELISA cortisol kits (K003-H5; cross reactivity with dexamethasone 18.8%, prednisolone (1-dehydrocortisol) 7.8%, corticosterone and cortisone 1.2%, other analytes < 0.1%; sensitivity 27.6pg/mL; limit of detection 45.4pg/mL). Plates were read at 450 nm on a Biotek Multi-Mode plate reader. LEEP conducted analytical validations of methods for adult female human saliva using tests of parallelism and accuracy for each analyte. Parallelism was done by serially diluting a sample pool, correcting the resulting concentrations at each dilution, and calculating the coefficient of variation (CV).^37^ The CV of corrected concentrations from serially diluted saliva was 11.65% for oxytocin (n=5 dilutions) and 8.34% for cortisol (n=10 dilutions). For accuracy, sample pools were spiked with 10% synthetic oxytocin or cortisol in assay buffer, at different concentrations along the standard curve. Recovery for salivary oxytocin was 119.99% ± 0.05 and recovery for salivary cortisol was 102.18% ± 2.31. All samples were assayed in duplicate, and all plates included high and low pool controls to assess inter- and intra-assay variation. Any samples with concentrations that were outside of the standard curve range were diluted or concentrated and re-assayed. Data were processed using BioTek Gen5 software to calculate analyte concentrations.

Using dried blood spots, ZRT Laboratory (Beaverton, OR) measured cortisol, estrone, estradiol, estriol, testosterone, progesterone, DHEAS, cortison, estrone-1-sulfate, pregnenlone sulfate, 17-hydroxyprogesterone, androstenedione, 7-keto DHEA, corticosterone, 11-dexycortisol, ethinyl estradiol, anastrozole, and letrozole as previously described with some modifications.^38^ Three 6mm diameter punches were taken from each dried blood spot specimen and rehydrated. Steroids were extracted with organic solvent in the presence of internal standard, purified by SPE, derivatized (estrogens only), and then analyzed by liquid chromatography-mass spectrometry (LC-MS/MS). LC-MS/MS analysis was performed using ultra-fast liquid chromatography system (Shimadzu Nexera XR) coupled to a triple quadrupole mass spectrometer (AB Sciez 5500) equipped with an electrospray ionization source. Separations were conducted by reversed-phase chromatography and the mass spectrometer was operated in multiple reaction monitoring mode. Data were processed using Sciex OS software to determine analyte concentrations. All samples were run with dried blood spot controls, generated in-house by spiking stripped serum with various analyte concentrations, mixing washed red blood cells, then spotting onto filter paper.

#### Parent Study Primary Outcome Variables

To assess postpartum risk for opioid misuse we included three outcomes. First, daily self-reports of opioid cravings (separately for opioid-containing prescription medications, heroin, and MOUD medications) and urge coping. Both cravings and urge coping were reported with a 100-point visual analog scale ranging from “not at all” to “severe” or “very well,” respectively. Second, daily self-reports of opioid use captured via daily surveys and staff-administered TimeLine FollowBack^39^ interview at each visit. Finally, we reviewed toxicology results and treatment adherence via treatment program medical records. We also compared OUD+ to OUD− on all predictor variables (maternal factors, caregiving factors, hormones) to identify levels/patterns specific to OUD+.

### Statistical Analyses

Study sample descriptors (e.g., sociodemographics, substance use) and outcomes were summarized with descriptives statistics. Group differences (i.e., OUD+ versus OUD−) were compared using Welch’s t-tests chi-squared tests, and Fisher’s Exact tests. All analyses were completed using open-source Python (3.10.11) libraries, Pandas 2.0.1 (NumFOCUS, Inc.), SciPy 1.13.0, and R Studio (2024.09.1, Build 394).

## Results

### Study Sample

There were 410 expressions of interest in study participation (Figure 1). Subsequently, 325 (79%) completed the eligibility survey, 113 (35%) completed the eligibility interview, and 104 (92%) met eligibility criteria. Of these, 71 (68%) completed baseline and enrolled. There was little to no evidence of selection bias during the enrollment process (Supplementary Table 2).

**Figure 1.**
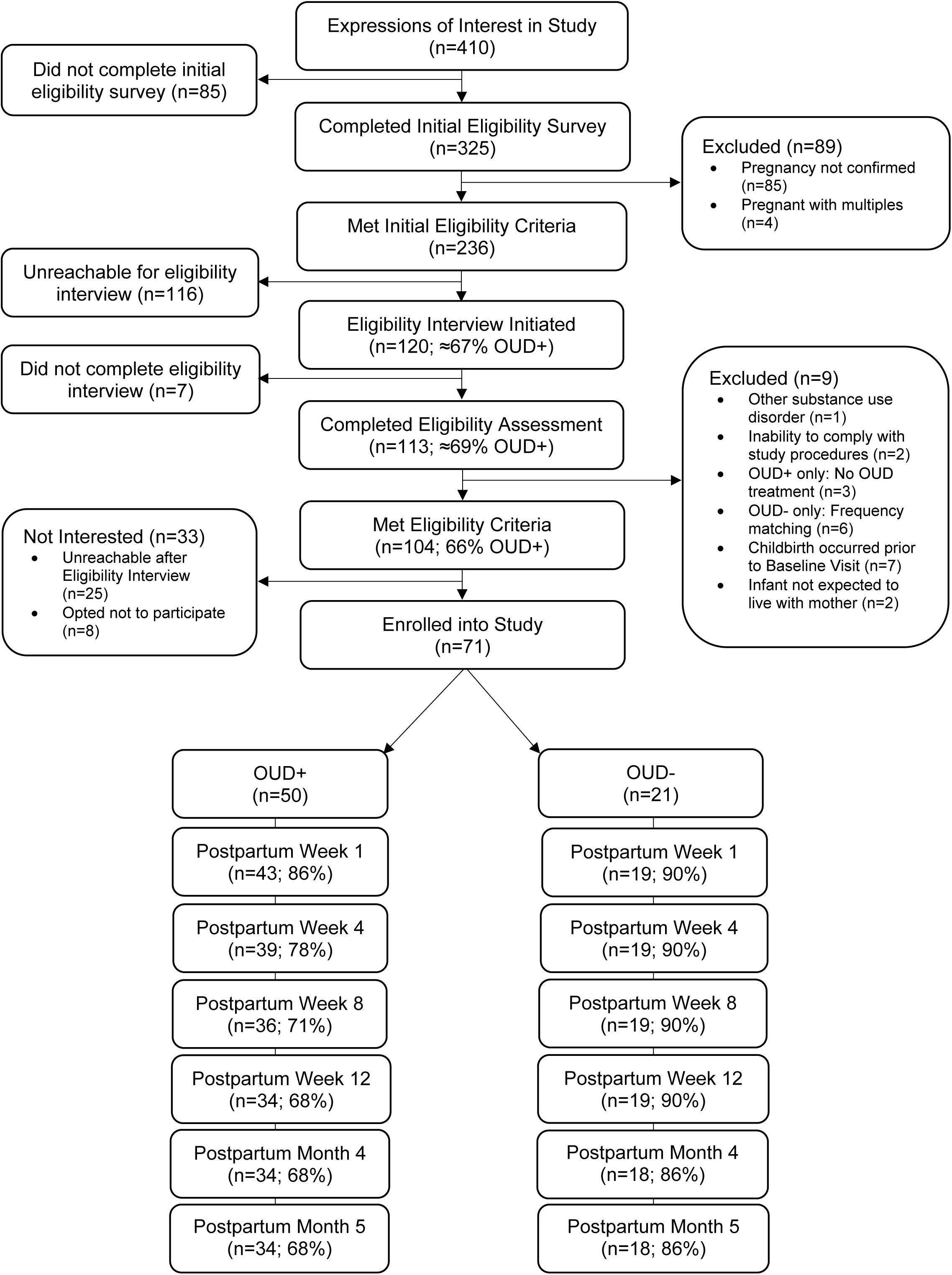
CONSORT Diagram: Study Flow, Timepoints and Participants.

The final sample included 50 (70%) OUD+ and 21 (30%) OUD− (Table 1). Participants were, on average, 29.4 (standard deviation [SD]: ±5.0) years old with 1.1 (SD±1.4) children living at home. Most were either Hispanic (48%) or non-Hispanic White (39%). Study groups were exchangeable on all sociodemographic variables except for gestational week prenatal care was initiated (OUD+: 9.8±5.2 versus OUD−: 7.1±3.7, respectively; p-value=0.01). OUD+ participants reported being in recovery for more than a year (42%), within the past year but before pregnancy (18%), or entered recovery during this pregnancy (40%). Most used MOUD (68%) in an outpatient setting (60%). Opioid-containing prescription medication was the most commonly endorsed opioid drug of choice (36%) followed by heroin (22%). Most reported use of narcotics (82%) and/or cigarettes or other nicotine-containing products (56%) during pregnancy. There was little to no evidence of selection bias between enrollment and the postpartum data collection period (Supplementary Table 3).

**Table 1.**
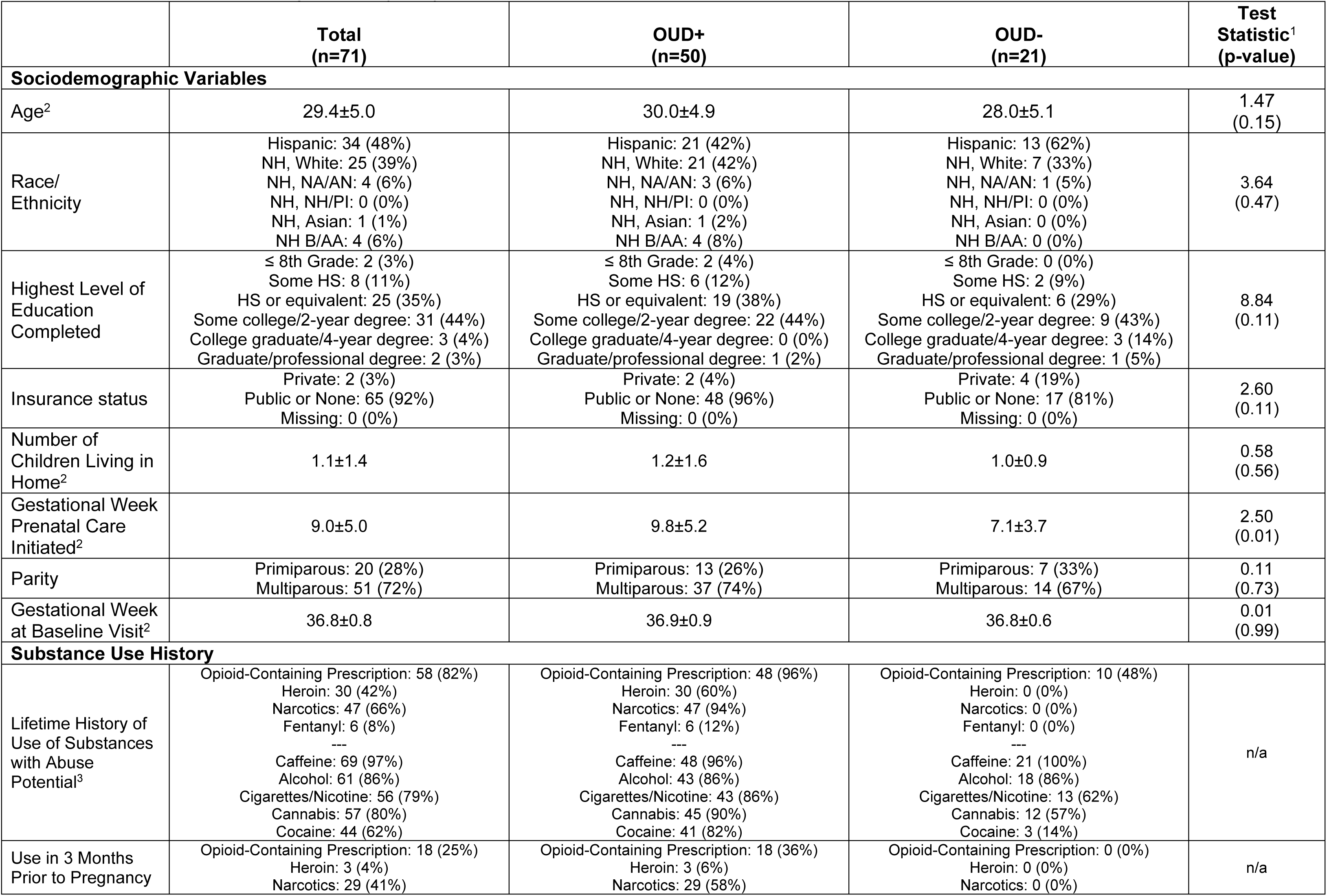
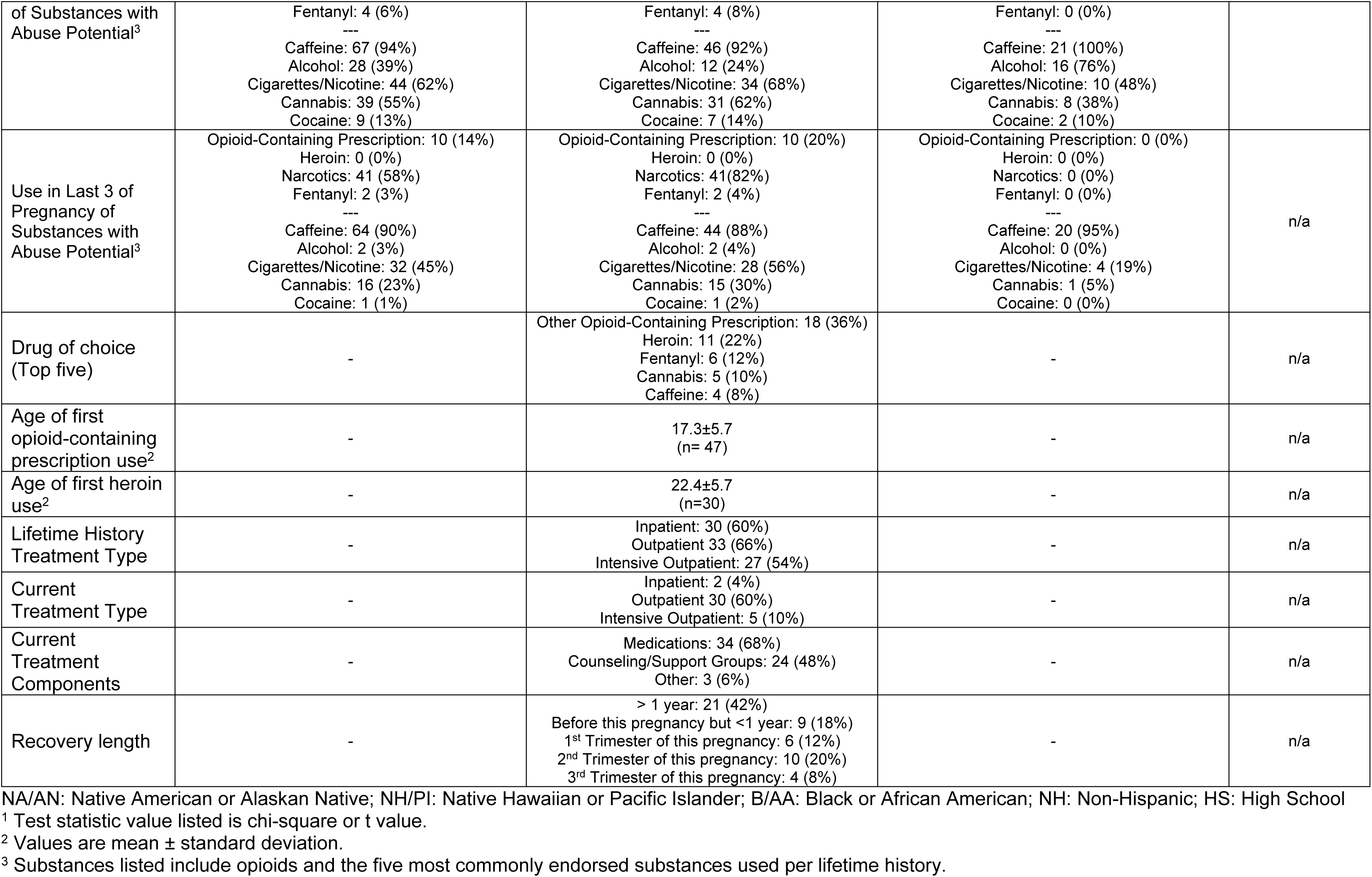
Description of Final Study Sample (n=71)

### Protocol Compliance

Nearly all participants (n=66, 93%) reported childbirth to staff (OUD+: 92%, OUD−:95%) and 62 (87%) completed at least one study visit post-childbirth (OUD+: 86%, OUD−: 90%). Visit completion rates post- baseline ranged from 83% at postpartum week 1 to 75% at postpartum month 5 (Figure 2; Supplementary Table 4), with lower rates in the OUD+ group and at later time points. Compliance with procedures followed similar patterns (lower in OUD+ and at later time points), with the highest overall compliance, on average, with the weekly surveys (78%) followed by interviews (74%), dried blood spots (69%), saliva samples (66%), and daily surveys (66%).

**Figure 2.**
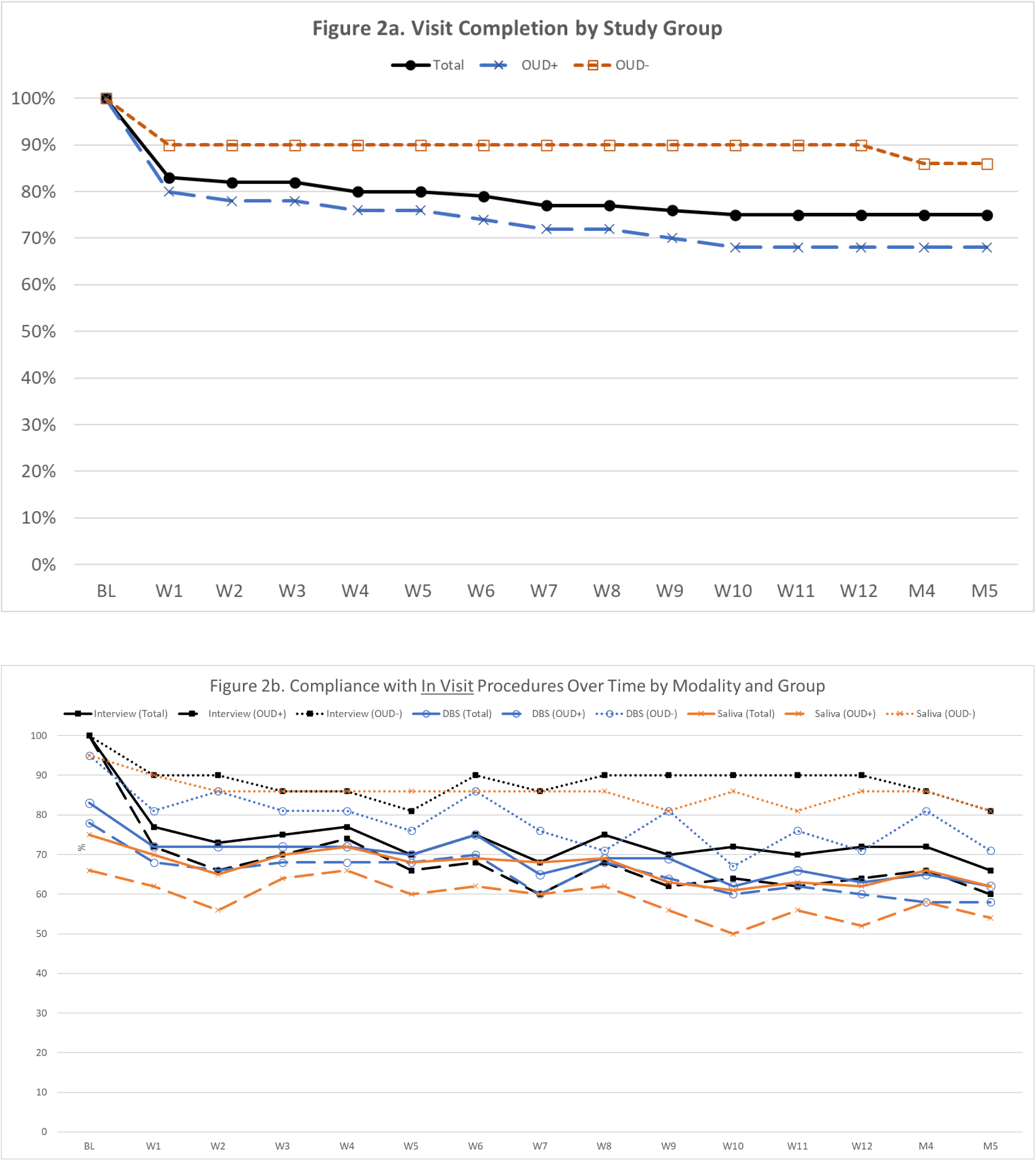
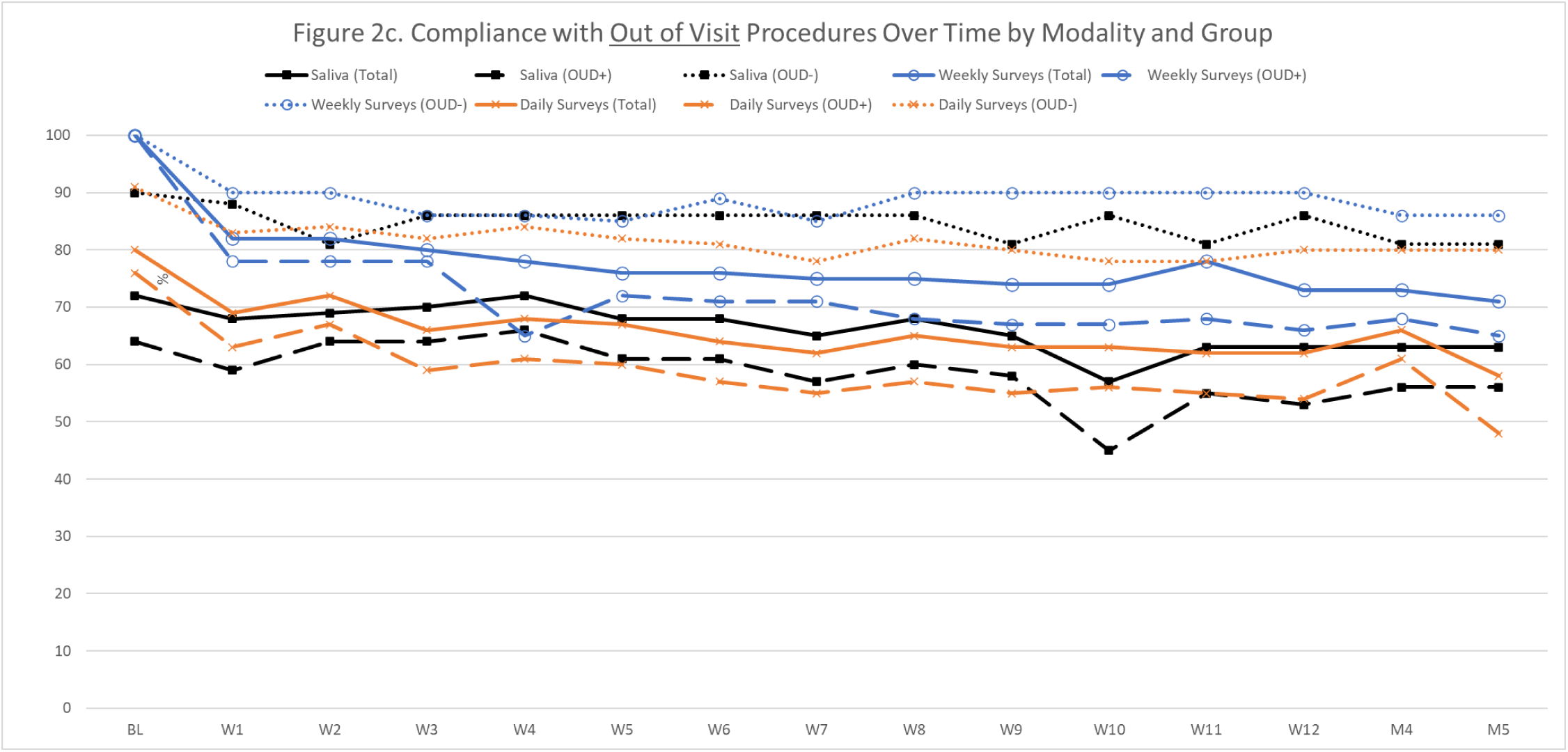
Compliance Rates by Study Group, Time Point, and Procedure.

### Study Satisfaction

Among the subgroup of participants who completed the study satisfaction surveys, results indicate that the study was well tolerated over time with no significant differences by study groups (Table 2).

## Discussion

We completed a prospective cohort study with individuals who do and do not have perinatal OUD following them with robust data collection procedures including daily surveys plus weekly face-to-face visits with collection of biological samples during the first 12 weeks postpartum with additional follow-up through postpartum month five. We enrolled 71 participants during late pregnancy and 52 (73%) completed through postpartum month five, with a higher completion rate in OUD− (86%) versus OUD+ (68%). Additionally, despite the challenges faced in caring for a newborn with our robust data collection protocol occurring concurrently, compliance with specific data collection modalities was moderately high in both groups (66-78%). The OUD+ group had lower compliance overall, which is to be expected given the additional demands on these participants (e.g., caring for a high-needs infant, daily MOUD clinic visits). Moreover, during the first year of data collection, participants faced additional challenges with the ongoing COVID-19 pandemic. Despite this, participants reported extremely high levels of satisfaction across study groups and time. Overall, execution of this novel protocol within a high-risk population during the challenging postpartum period has yielded ample opportunity to explore novel predictors of OUD recovery-related outcomes. These critically important data will ultimately inform new relapse prevention interventions tailored to postpartum-specific needs and opportunities.

The forthcoming primary goal of this prospective cohort study is to examine the potential utility of hormones and/or infant caregiving activities to support postpartum OUD recovery. While there is growing evidence that hormones (e.g., progesterone, estrogen, oxytocin) influence drug-taking behaviors, nearly all research has examined the effect of a single hormone in isolation at one or two timepoints.^15–17^ Indeed, emerging research demonstrates relative levels and temporal patterns of hormones may be more important than a single hormone value at one time point. For example, changes in the ratio of progesterone to estradiol across two time points is more predictive of smoking behavior than either hormone alone at a single time point.^40^ Given that hormones: (a) are continuously in flux, especially during the perinatal period, and variations occur as a result of many factors, including specific infant caregiving activities, and (b) have a significant effect on several aspects of neurobiology, including drug reward and stress responses, assessing the effects of one or two hormones at one or two time points is insufficient. We will examine the role of hormones and infant caregiving practices on OUD recovery-related outcomes with novel analytical approaches to model complex variable interactions both concurrently and across time. This will ultimately yield enhanced accuracy and generalizability of observations.

Beyond our primary goals, these data provide ample opportunities to explore novel topics in perinatal substance misuse and beyond. For example, we conducted comprehensive interviews of lifetime trauma exposure and resilience, as well as birth and breastfeeding experiences. This allows for mixed method approaches to expand the understanding of these experiences prospectively during the perinatal period. Additionally, daily our participants reported on their mood, affect, and stress, as well as their craving for and use of other substances (e.g., alcohol, cannabis, nicotine/tobacco). Thus, we can explore relationships between these variables, as well as in relation to baseline characteristics (e.g., ethnicity, parity) and concurrent behaviors and activities (e.g., sleep, involvement of others). Indeed, we are currently exploring the role of loneliness, isolation, and social support in OUD−related outcomes within this dataset (R21DA058364; Allen & Linde-Krieger). Overall, these data offer substantial opportunities to uncover novel relationships, increase our understanding risk/protective factors of maladaptive behaviors during the sensitive perinatal period, and ultimately inform new approaches to enhance the health and well-being of mothers, infants, and families.

Like every study, there are limitations. First, generalizability is limited given this data was collected in a single US state and the robust protocol may have been too burdensome for some individuals. Similarly, our eligibility criteria restricted our OUD+ group to those who were in treatment for recovery and expected to live with their infant. However, our findings here indicate little to no selection bias. We also have a fair amount of missing data, as is common in prospective studies. These data may not be missing at random (e.g., those with a high-needs newborn may be less likely to complete data collection and also may be more likely to be at risk for return to opioid misuse). It is possible that this will introduce bias and error into our observations. Despite these limitations, there are also substantial strengths including an ability to temporally evaluate relationships, assessment of substance use with multiple sources (e.g., self-report, medical records), and numerous validated measures with rigorous, gold-standard approaches.

Overall, this prospective cohort study of those with and without OUD during the perinatal period resulted in high retention and compliance, as well as exceptionally high participant-reported study satisfaction. This indicates that this robust protocol with weekly biological sample collection and daily report of subjective experiences is feasible in the early postpartum, even among those who may face additional challenges during this time. These multidimensional data have high potential to provide new insights into the challenges and opportunities of the perinatal period. Perhaps most promising, this study will uncover complex variable interactions across time informing relapse prevention efforts to promote positive OUD recovery and overall well-being during the traditionally high-risk postpartum period.

## Data Availability

All data produced in the present study are available upon reasonable request to the authors.

## Acknowledgements

We extend our thanks to Drs. Mustafa al’Absi, Ann Bigelow, Ariadna Forray, Bob Handa, and Toni Ziegler for their input during the development of the initial data collection protocol. We also thank Emily Crose, Jeri Garfield, Rosalie Gordon, Kyndall Kleinman, Kristina Medvescek, McKenna Nelson, and Preeti Sahota for their participation in database development, recruitment, and data collection.

## Ethical approval

All procedures performed in studies involving human participants were in accordance with the ethical standards of the institutional and/or national research committee and with the 1964 Helsinki declaration and its later amendments or comparable ethical standards.

## Author Agreement

All authors have provided substantial contributions to this work, participated in the writing and revising of this manuscript, seen and approved the final version of this manuscript, and agree to be accountable for all aspects of this work. This manuscript is not being considered elsewhere for publication and has not already been published.

