## Supplementary Table 1 for "Hormones and Infant Caregiving in Postpartum Opioid Use Disorder Recovery: Compliance and Satisfaction with a Prospective Cohort Study Protocol Designed to Identify Novel Support Targets"

**Supplementary Table 1. Overview of Study Measures**

| | Timepoint of Collection | Mode of Collection | Location of Collection | Reference and/or Supplement Document |
|---|---|---|---|---|
| **Background Variables** | | | | |
| Sociodemographics | Enrollment and Baseline | Self-Administered Survey and Interview with Staff | At home and during visit | Supplementary Document 3 |
| Medical History | Enrollment and Baseline | Self-Administered Survey and Interview with Staff | At home and during visit | Supplementary Document 3 |
| Substance Use History | Enrollment and Baseline | Self-Administered Survey and Interview with Staff | At home and during visit | Supplementary Document 3 |
| Stressful Life Events | Baseline | Audio-Recorded Interview with Staff | During visit | Newton et al,[32] Supplementary Document 1 |
| Adverse Childhood Experiences | Baseline | Audio-Recorded Interview with Staff | During visit | Felitti et al,[33] Supplementary Document 1 |
| Modified Early Trauma Inventory | Baseline | Audio-Recorded Interview with Staff | During visit | Bremner et al,[34,35] Supplementary Document 1 |
| Resilience Scale | Baseline | Audio-Recorded Interview with Staff | During visit | Wagnild et al,[36] Supplementary Document 1 |
| Birth Interview | PW1 | Audio-Recorded Interview with Staff | During visit | Supplementary Document 2 |
| **Maternal Variables** | | | | |
| Breastfeeding/Lactation | Daily PW 1-4, Weekly PW 5-12, PM 4, PM 5 | Self-Administered Survey and Audio-Recorded Interview with Staff | At home and during visit | Supplementary Document 2 and 3 (Daily Assessment) |
| Vaginal Bleeding | Daily PW 5-12, PM 4, PM 5 | Self-Administered Survey | At home | Supplementary Document 3 (Daily Assessment) |
| Modified Brief Pain Inventory, Pregnancy and Postpartum Pain Inventory | Baseline, Weekly PW 1-4 | Self-Administered Survey | At home | Cleeland et al,[37] Supplementary Document 3 |
| Edinburgh Postnatal Depression Scale | Baseline, Weekly PW 1-12, PM 4, PM 5 | Self-Administered Survey | At home | Cox et al[38] |
| Modified Postpartum Stressor Scale | Baseline, Weekly PW 1-12, PM 4, PM 5 | Self-Administered Survey | At home | Park et al,[39] Supplementary Document 3 |
| Depression Anxiety Stress Scale | Baseline, Weekly PW 1-12, PM 4, PM 5 | Self-Administered Survey | At home | Osman et al[40] |
| Postpartum Bonding Questionnaire | Baseline, Weekly PW 1-12, PM 4, PM 5 | Self-Administered Survey | At home | Brockington et al [41] |

| | | | | |
|---|---|---|---|---|
| Karitane Parenting Confidence Scale | Baseline, Weekly PW 1-12, PM 4, PM 5 | Self-Administered Survey | At home | Črnčec et al [42] |
| Modified Epworth Sleepiness Scale | Baseline, Weekly PW 1-12, PM 4, PM 5 | Self-Administered Survey | At home | Johns et al,[43] Supplementary Document 3 |
| Modified Pittsburgh Sleep Quality Index | Baseline, Monthly PW 1-12, PM 4, PM 5 | Self-Administered Survey | At home | Buysse et al, [44] Supplementary Document 3 |
| UCLA Loneliness Scale-Revised | Baseline, Monthly PW 1-12, PM 4, PM 5 | Self-Administered Survey | At home | Russell et al[45] |
| MOS Social Support Scale | Baseline, Monthly PW 1-12, PM 4, PM 5 | Self-Administered Survey | At home | Sherbourne et al[46] |
| Barkin Index of Maternal Functioning | Baseline, Monthly PW 1-12, PM 4, PM 5 | Self-Administered Survey | At home | Barkin et al[47] |
| Reflective Functioning Questionnaire | Baseline | Self-Administered Survey | At home | Fonagy et al[50] |
| Parental Reflective Functioning Questionnaire | PW 12, PM 4, PM 5 | Self-Administered Survey | At home | Luyten et al[49] |
| **Caregiving Variables** | | | | |
| Time Spent with Infant | Daily and Weekly PW 1-12, PM 4, PM 5 | Self-Administered Survey and Interview with Staff | At home and during visit | Supplementary Document 3 (Daily Assessment) |
| Subjective Response to Parenting | Daily PW 1-12, PM 4, PM 5 | Self-Administered Survey | At home | Supplementary Document 3 (Daily Assessment) |
| Infant Behavior Questionnaire | Weekly PW 1-12, PM 4, PM 5 | Self-Administered Survey | At home | Gradstein et al[51] |
| Modified Brief Infant Sleep Questionnaire | Weekly PW 1-12, PM 4, PM 5 | Self-Administered Survey | At home | Sadeh et al[52] Supplementary Document 3 |
| Parenting Sense of Competency Scale | Monthly PW 1-12, PM 4, PM 5 | Self-Administered Survey | At home | Ohan et al[48] |
| Modified Father and Other Involvement Survey | Monthly PW 1-12, PM 4, PM 5 | Self-Administered Survey | At home | Wood et al[53] Supplementary Document 3 |
| **Hormones** | | | | |
| Oxytocin | Baseline, Weekly PW 1-12, PM 4, PM 5 | Saliva Sample | During visit | - |
| Cortisol | Baseline, Weekly PW 1-12, PM 4, PM 5 | Saliva Sample | At home (8pm and 30 minutes after waking) | - |
| Cortisol, Estrone, Estradiol, Estriol, Testosterone, Progesterone, DHEAS, Cortison, Estrone-1-sulfate, Pregnenlone Sulfate, 17- | Baseline, Weekly PW 1-12, PM 4, PM 5 | Dried Blood Spots | During visit | - |

| | | | | |
|---|---|---|---|---|
| hydroxyprogesterone, Androstenedione, 7-Keto DHEA, Corticosterone, 11-Dexycortisol, Ethinyl Estradiol, Anastrozole, Letrozole | | | | |
| **Outcome Variables** | | | | |
| Craving | Baseline, Daily PW 1-12, PM 4, PM 5 | Self-Administered Survey | At home | Supplementary Document 3 (Daily Assessment) |
| Urge Coping | Baseline, Daily PW 1-12, PM 4, PM 5 | Self-Administered Survey | At home | Supplementary Document 3 (Daily Assessment) |
| Use (Prospective) | Baseline, Daily PW 1-12, PM 4, PM 5 | Self-Administered Survey | At home | Supplementary Document 3 (Daily Assessment) |
| Use (Retrospective via TimeLine FollowBack) | Baseline, Weekly PW 1-12, PM 4, PM 5 | Interview with Staff | During visit | Sobell et al [56] Supplementary Document 3 (Daily Assessment) |
| Toxicology Results | Childbirth to One Year Postpartum | Medical Record Chart Review | n/a | - |
| Treatment Program Adherence | Childbirth to One Year Postpartum | Medical Record Chart Review | n/a | - |
