## Supplementary Table 2 for "Hormones and Infant Caregiving in Postpartum Opioid Use Disorder Recovery: Compliance and Satisfaction with a Prospective Cohort Study Protocol Designed to Identify Novel Support Targets"

**Supplementary Table 2. Description of Study Sample – Completed Phone Screening Interview (n=113)**

| | Total (n=113) | OUD+ (n=74) | OUD- (n=39) | Test Statistic[1] (p-value) |
|---|---|---|---|---|
| Age (mean ± standard deviation) | 28.6±5.2 | 28.8±5.2 | 28.2±5.1 | 0.61 (0.54) |
| Race/Ethnicity | Hispanic: 59 (52%)<br>NH, White: 43 (38%)<br>NH, NA/AN: 4 (4%)<br>NH, NH/PI: 2 (2%)<br>NH, Asian: 1 (1%)<br>NH, B/AA: 4 (4%) | Hispanic: 36 (49%)<br>NH, White: 30 (41%)<br>NH, NA/AN: 3 (4%)<br>NH, NH/PI: 0 (0%)<br>NH, Asian: 1 (1%)<br>NH, B/AA: 4 (5%) | Hispanic: 23 (59%)<br>NH, White: 13 (33%)<br>NH, NA/AN: 1 (3%)<br>NH, NH/PI: 2 (5%)<br>NH, Asian: 0 (0%)<br>NH, B/AA: 0 (0%) | 7.46 (0.19) |
| Highest Level of Education Completed | ≤ 8th Grade: 2 (2%)<br>Some HS: 18 (16%)<br>HS or equivalent: 42 (37%)<br>Some college/2-year degree: 42 (37%)<br>College graduate/4-year degree: 4 (4%)<br>Graduate/professional degree: 5 (4%) | ≤ 8th Grade: 2 (2%)<br>Some HS: 14 (19%)<br>HS or equivalent: 28 (38%)<br>Some college/2-year degree: 29 (39%)<br>College graduate/4-year degree: 0 (0%)<br>Graduate/professional degree: 1 (1%) | ≤ 8th Grade: 0 (0%)<br>Some HS: 4 (10%)<br>HS or equivalent: 14 (36%)<br>Some college/2-year degree: 13 (33%)<br>College graduate/4-year degree: 4 (10%)<br>Graduate/professional degree: 4 (10%) | 14.68 (0.01) |
| Insurance Status | Private: 11 (10%)<br>Public or None: 99 (88%)<br>Missing: 3 (3%) | Private: 3 (4%)<br>Public or None: 70 (95%)<br>Missing: 1 (1%) | Private: 8 (21%)<br>Public or None: 29 (74%)<br>Missing: 2 (5%) | 12.62, (<0.01) |

NA/AN: Native American or Alaskan Native; NH/PI: Native Hawaiian or Pacific Islander; B/AA: Black or African American; NH: Non-Hispanic; HS: High School

[1] Test statistic value listed is chi-square or t value.
