## Supplementary Table 3 for "Hormones and Infant Caregiving in Postpartum Opioid Use Disorder Recovery: Compliance and Satisfaction with a Prospective Cohort Study Protocol Designed to Identify Novel Support Targets"

**Supplementary Table 3. Description of Study Sample – Completed at least 1 Postpartum Visit (n=62)**

| | Total (n=62) | OUD+ (n=43) | OUD- (n=19) | Test Statistic[1] (p-value) |
|---|---|---|---|---|
| **Sociodemographic Variables** | | | | |
| Age[2] | 29.5±5.1 | 30.1±5.0 | 28.0±5.2 | 1.47 (0.15) |
| Race/Ethnicity | Hispanic: 32 (52%)<br>NH, White: 25 (40%)<br>NH, NA/AN: 3 (5%)<br>NH, NH/PI: 0 (0%)<br>NH, Asian: 1 (2%)<br>NH B/AA: 1 (2%) | Hispanic: 21 (49%)<br>NH, White: 18 (42%)<br>NH, NA/AN: 2 (5%)<br>NH, NH/PI: 0 (0%)<br>NH, Asian: 1 (2%)<br>NH B/AA: 1 (2%) | Hispanic: 11 (58%)<br>NH, White: 7 (37%)<br>NH, NA/AN: 1 (5%)<br>NH, NH/PI: 0 (0%)<br>NH, Asian: 0 (0%)<br>NH B/AA: 0 (0%) | 1.18 (0.88) |
| Highest Level of Education Completed | ≤ 8th Grade: 2 (3%)<br>Some HS: 8 (13%)<br>HS or equivalent: 22 (35%)<br>Some college/2-year degree: 26 (42%)<br>College graduate/4-year degree: 2 (3%)<br>Graduate/professional degree: 2 (3%) | ≤ 8th Grade: 2 (4%)<br>Some HS: 6 (14%)<br>HS or equivalent: 17 (39%)<br>Some college/2-year degree: 17 (39%)<br>College graduate/4-year degree: 0 (0%)<br>Graduate/professional degree: 1 (2%) | ≤ 8th Grade: 0 (0%)<br>Some HS: 2 (10%)<br>HS or equivalent: 5 (26%)<br>Some college/2-year degree: 9 (47%)<br>College graduate/4-year degree: 2 (10%)<br>Graduate/professional degree: 1 (5%) | 6.72 (0.24) |
| Insurance Status | Private: 5 (8%)<br>Public or None: 57 (92%)<br>Missing: 0 (0%) | Private: 1 (2%)<br>Public or None: 42 (97%)<br>Missing: 0 (0%) | Private: 4 (21%)<br>Public or None: 15 (79%)<br>Missing: 0 (0%) | 3.96 (0.05) |
| Number of Children Living in Home[2] | 1.1±1.5 | 1.2±1.7 | 0.9±0.8 | 0.87 (0.38) |
| Gestational Week Prenatal Care Initiated[2] | 8.9±5.1 | 9.7±5.4 | 7.2±3.8 | 2.03 (0.05) |
| Parity | Primiparous: 18 (29%)<br>Multiparous:44 (71%) | Primiparous: 11 (25%)<br>Multiparous: 32 (74%) | Primiparous: 7 (37%)<br>Multiparous: 12 (63%) | 0.36 (0.55) |
| Gestational Week at Baseline Visit[2] | 36.8±0.8 | 36.8±0.8 | 36.9±0.6 | 0.29 (0.77) |
| Gestational Week at time of delivery[2] | 39.2±1.1 | 39.3±1.7 | 39.2±0.8 | 0.22 (0.82) |
| Mother/Infant Discharged from Hospital at Same Time | 44 (71%) | 27 (63%) | 17 (89%) | 3.35 (0.07) |
| **Substance Use History** | | | | |
| Lifetime History of Use of | Opioid-Containing Prescription: 51 (82%)<br>Heroin: 26 (42%)<br>Narcotics: 41 (66%) | Opioid-Containing Prescription: 42 (98%)<br>Heroin: 26 (60%)<br>Narcotics: 41 (95%) | Opioid-Containing Prescription: 9 (47%)<br>Heroin: 0 (0%)<br>Narcotics: 0 (0%) | n/a |

| | | | | |
|---|---|---|---|---|
| Substances with Abuse Potential[3] | Fentanyl: 5 (8%)<br>----<br>Caffeine: 60 (97%)<br>Alcohol: 54 (87%)<br>Cigarettes/Nicotine: 50 (81%)<br>Cannabis: 49 (79%)<br>Cocaine: 37 (60%) | Fentanyl: 5 (12%)<br>----<br>Caffeine: 41 (95%)<br>Alcohol: 38 (88%)<br>Cigarettes/Nicotine: 39 (91%)<br>Cannabis: 39 (91%)<br>Cocaine: 36 (84%) | Fentanyl: 0 (0%)<br>---<br>Caffeine: 19 (100%)<br>Alcohol: 16 (84%)<br>Cigarettes/Nicotine: 11 (58%)<br>Cannabis: 10 (53%)<br>Cocaine: 1 (5%) |  |
| Use in 3 Months Prior to Pregnancy of Substances with Abuse Potential[3] | Opioid-Containing Prescription: 13 (21%)<br>Heroin: 1 (2%)<br>Narcotics: 25 (40%)<br>Fentanyl: 4 (6%)<br>---<br>Caffeine: 58 (94%)<br>Alcohol: 25 (40%)<br>Cigarettes/Nicotine: 38 (61%)<br>Cannabis: 32 (52%)<br>Cocaine: 4 (6%) | Opioid-Containing Prescription: 13 (30%)<br>Heroin: 1 (2%)<br>Narcotics: 25 (58%)<br>Fentanyl: 4 (9%)<br>---<br>Caffeine: 39 (91%)<br>Alcohol: 10 (23%)<br>Cigarettes/Nicotine: 30 (70%)<br>Cannabis: 26 (60%)<br>Cocaine: 4 (9%) | Opioid-Containing Prescription: 0 (0%)<br>Heroin: 0 (0%)<br>Narcotics: 0 (0%)<br>Fentanyl: 0 (0%)<br>---<br>Caffeine: 19 (100%)<br>Alcohol: 15 (79%)<br>Cigarettes/Nicotine: 8 (42%)<br>Cannabis: 6 (32%)<br>Cocaine: 0 (0%) | n/a |
| | Opioid-Containing Prescription: 7 (11%)<br>Heroin: 0 (0%)<br>Narcotics: 35 (56%)<br>Fentanyl: 1 (2%)<br>---<br>Caffeine: 55 (89%)<br>Alcohol: 2 (3%)<br>Cigarettes/Nicotine: 28 (45%)<br>Cannabis: 13 (21%)<br>Cocaine: 1 (2%) | Opioid-Containing Prescription: 7 (16%)<br>Heroin: 0 (0%)<br>Narcotics: 35 (81%)<br>Fentanyl: 1 (2%)<br>---<br>Caffeine: 37 (86%)<br>Alcohol: 2 (5%)<br>Cigarettes/Nicotine: 25 (58%)<br>Cannabis: 13 (30%)<br>Cocaine: 1 (2%) | Opioid-Containing Prescription: 0 (0%)<br>Heroin: 0 (0%)<br>Narcotics: 0 (0%)<br>Fentanyl: 0 (0%)<br>---<br>Caffeine: 18 (95%)<br>Alcohol: 0 (0%)<br>Cigarettes/Nicotine: 3 (16%)<br>Cannabis: 0 (0%)<br>Cocaine: 0 (0%) | n/a |
| Drug of choice (Top five) | - | Opioid-Containing Prescription: 16 (37%)<br>Heroin: 11 (26%)<br>Fentanyl: 6 (14%)<br>Cannabis: 3 (7%)<br>Caffeine: 3 (7%) | - | n/a |
| Age of first opioid-containing prescription use[2] | - | 17.5±5.9<br>(n=41) | - | n/a |
| Age of first heroin use[2] | - | 22.6±5.6<br>(n=26) | - | n/a |
| Lifetime History Treatment Type | - | Inpatient: 27 (63%)<br>Outpatient 26 (61%)<br>Intensive Outpatient: 23 (54%) | - | n/a |
| Current Treatment Type | - | Inpatient: 2 (5%)<br>Outpatient 25 (58%)<br>Intensive Outpatient: 5 (12%) | - | n/a |

| | | | | |
|---|---|---|---|---|
| Current Treatment Components | - | Medication: 34 (79%)<br>Counseling/Support Groups: 23 (54%)<br>Other: 2 (5%) | - | n/a |
| Length of Recovery | - | > 1 year: 19 (44%)<br>Before this pregnancy but <1 year: 8 (19%)<br>1st Trimester of this pregnancy: 5 (12%)<br>2nd Trimester of this pregnancy: 8 (19%)<br>3rd Trimester of this pregnancy: 3 (7%) | - | n/a |

NA/AN: Native American or Alaskan Native; NH/PI: Native Hawaiian or Pacific Islander; B/AA: Black or African American; NH: Non-Hispanic; HS: High School

[1] Test statistic value listed is chi-square or t value.

[2] Values are mean ± standard deviation.

[3] Substances listed include opioids and the five most commonly endorsed substances used per lifetime history.
