## Supplementary Table 4 for "Hormones and Infant Caregiving in Postpartum Opioid Use Disorder Recovery: Compliance and Satisfaction with a Prospective Cohort Study Protocol Designed to Identify Novel Support Targets"

**Table 2. Protocol Completion by Time Point, Modality, and Group**

| | Total (n=71) | OUD+ (n=50) | OUD- (n=21) | t-value, p-value |
|---|---|---|---|---|
| **Baseline Visit Completed** | **71 (100%)** | **50 (100%)** | **21 (100%)** | **-** |
| Interviews (During Visit) | 71 (100%) | 50 (100%) | 21 (100%) | - |
| Dried Blood Spots (During Visit) | 59 (83%) | 39 (78%) | 20 (95%) | 2.02, 0.15 |
| Saliva Samples (During Visit) | 53 (75%) | 33 (66%) | 20 (95%) | 5.22, 0.02 |
| Saliva Samples (Outside Visit) | 102 (72%) | 64 (64%) | 38 (90%) | 8.98, <0.01 |
| Daily Surveys (Outside Visit) * | 380 (80%) | 254 (76%) | 126 (91%) | 13.0, <0.01 |
| Weekly Surveys (Outside Visit) | 639 (100%) | 450 (100%) | 189 (100%) | - |
| **Week 1 Visit Completed** | **59 (83%)** | **40 (80%)** | **19 (90%)** | **0.53, 0.47** |
| Interviews (During Visit) | 55 (77%) | 36 (72%) | 19 (90%) | 1.93, 0.16 |
| Dried Blood Spots (During Visit) | 51 (72%) | 34 (68%) | 17 (81%) | 0.67, 0.41 |
| Saliva Samples (During Visit) | 50 (70%) | 31 (62%) | 19 (90%) | 4.47, 0.03 |
| Saliva Samples (Outside Visit) | 96 (68%) | 59 (59%) | 37 (88%) | 10.14, <0.01 |
| Daily Surveys (Outside Visit) | 344 (69%) | 222 (63%) | 122 (83%) | 17.69, <0.01 |
| Weekly Surveys (Outside Visit) | 523 (82%) | 352 (78%) | 171 (90%) | 12.64, <0.01 |
| **Week 2 Visit Completed** | **58 (82%)** | **39 (78%)** | **19 (90%)** | **0.82, 0.36** |
| Interviews (During Visit) | 52 (73%) | 33 (66%) | 19 (90%) | 3.36, 0.07 |
| Dried Blood Spots (During Visit) | 51 (72%) | 33 (66%) | 18 (86%) | 1.95, 0.16 |
| Saliva Samples (During Visit) | 46 (65%) | 28 (56%) | 18 (86%) | 4.49, 0.03 |
| Saliva Samples (Outside Visit) | 98 (69%) | 64 (64%) | 34 (81%) | 3.22, 0.07 |
| Daily Surveys (Outside Visit) | 357 (72%) | 233 (67%) | 124 (84%) | 15.31, <0.01 |
| Weekly Surveys (Outside Visit) | 523 (82%) | 353 (78%) | 170 (90%) | 11.09, <0.01 |
| **Week 3 Visit Completed** | **58 (82%)** | **39 (78%)** | **19 (90%)** | **0.82, 0.36** |
| Interviews (During Visit) | 53 (75%) | 35 (70%) | 18 (86%) | 1.19, 0.27 |
| Dried Blood Spots (During Visit) | 51 (72%) | 34 (68%) | 17 (81%) | 0.67, 0.41 |
| Saliva Samples (During Visit) | 50 (70%) | 32 (64%) | 18 (86%) | 2.39, 0.12 |
| Saliva Samples (Outside Visit) | 100 (70%) | 64 (64%) | 36 (86%) | 5.69, 0.02 |
| Daily Surveys (Outside Visit) | 327 (66%) | 207 (59%) | 120 (82%) | 22.28, <0.01 |
| Weekly Surveys (Outside Visit) | 512 (80%) | 350 (78%) | 162 (86%) | 4.78, 0.03 |
| **Week 4 Visit Completed** | **57 (80%)** | **38 (76%)** | **19 (90%)** | **1.15, 0.28** |
| Interviews (During Visit) | 55 (77%) | 37 (74%) | 18 (86%) | 0.59, 0.44 |
| Dried Blood Spots (During Visit) | 51 (72%) | 34 (68%) | 17 (81%) | 0.67, 0.41 |
| Saliva Samples (During Visit) | 51 (72%) | 33 (66%) | 18 (86%) | 1.95, 0.16 |
| Saliva Samples (Outside Visit) | 102 (72%) | 66 (66%) | 36 (86%) | 4.75, 0.03 |
| Daily Surveys (Outside Visit) | 338 (68%) | 215 (61%) | 123 (84%) | 22.53, <0.01 |
| Weekly Surveys (Outside Visit) | 884 (78%) | 586 (75%) | 288 (86%) | 20.02, <0.01 |
| **Week 5 Visit Completed** | **57 (80%)** | **38 (76%)** | **19 (90%)** | **1.15, 0.28** |
| Interviews (During Visit) | 50 (70%) | 33 (66%) | 17 (81%) | 0.95, 0.33 |
| Dried Blood Spots (During Visit) | 50 (70%) | 34 (68%) | 16 (76%) | 0.16, 0.68 |
| Saliva Samples (During Visit) | 48 (68%) | 30 (60%) | 18 (86%) | 3.37, 0.06 |
| Saliva Samples (Outside Visit) | 97 (68%) | 61 (61%) | 36 (86%) | 7.24, <0.01 |
| Daily Surveys (Outside Visit) | 331 (67%) | 210 (60%) | 121 (82%) | 22.18, <0.01 |
| Weekly Surveys (Outside Visit) | 483 (76%) | 322 (72%) | 161 (85%) | 12.67, <0.01 |
| **Week 6 Visit Completed** | **56 (79%)** | **37 (74%)** | **19 (90%)** | **1.52, 0.22** |
| Interviews (During Visit) | 53 (75%) | 34 (68%) | 19 (90%) | 2.85, 0.09 |
| Dried Blood Spots (During Visit) | 53 (75%) | 35 (70%) | 18 (86%) | 1.19, 0.27 |
| Saliva Samples (During Visit) | 49 (69%) | 31 (62%) | 18 (86%) | 2.86, 0.09 |
| Saliva Samples (Outside Visit) | 97 (68%) | 61 (61%) | 36 (86%) | 7.24, **<0.01** |
| Daily Surveys (Outside Visit) | 318 (64%) | 199 (57%) | 119 (81%) | 25.04, **<0.01** |
| Weekly Surveys (Outside Visit) | 486 (76%) | 318 (71%) | 168 (89%) | 23.28, **<0.01** |
| **Week 7 Visit Completed** | **55 (77%)** | **36 (72%)** | **19 (90%)** | **1.93, 0.16** |
| Interviews (During Visit) | 48 (68%) | 30 (60%) | 18 (86%) | 3.37, 0.06 |
| Dried Blood Spots (During Visit) | 46 (65%) | 30 (60%) | 16 (76%) | 1.06, 0.30 |
| Saliva Samples (During Visit) | 48 (68%) | 30 (60%) | 18 (86%) | 3.37, 0.06 |
| Saliva Samples (Outside Visit) | 93 (65%) | 57 (57%) | 36 (86%) | 9.56, <0.01 |
| Daily Surveys (Outside Visit) | 308 (62%) | 193 (55%) | 115 (78%) | 22.45, <0.01 |
| Weekly Surveys (Outside Visit) | 479 (75%) | 318 (71%) | 161 (85%) | 14.18, <0.01 |

| | | | | |
|---|---|---|---|---|
| **Week 8 Visit Completed** | **55 (77%)** | **36 (72%)** | **19 (90%)** | **1.93, 0.16** |
| Interviews (During Visit) | 53 (75%) | 34 (68%) | 19 (90%) | 2.85, 0.09 |
| Dried Blood Spots (During Visit) | 49 (69%) | 34 (68%) | 15 (71%) | 0.00, 1.00 |
| Saliva Samples (During Visit) | 49 (69%) | 31 (62%) | 18 (86%) | 2.86, 0.09 |
| Saliva Samples (Outside Visit) | 96 (68%) | 60 (60%) | 36 (86%) | 7.79, <0.01 |
| Daily Surveys (Outside Visit) | 322 (65%) | 201 (57%) | 121 (82%) | 27.02, <0.01 |
| Weekly Surveys (Outside Visit) | 847 (75%) | 546 (68%) | 301 (90%) | 55.65, <0.01 |
| **Week 9 Visit Completed** | **54 (76%)** | **35 (70%)** | **19 (90%)** | **2.39, 0.12** |
| Interviews (During Visit) | 50 (70%) | 31 (62%) | 19 (90%) | 4.47, 0.03 |
| Dried Blood Spots (During Visit) | 49 (69%) | 32 (64%) | 17 (81%) | 1.27, 0.26 |
| Saliva Samples (During Visit) | 45 (63%) | 28 (56%) | 17 (81%) | 2.96, 0.08 |
| Saliva Samples (Outside Visit) | 92 (65%) | 58 (58%) | 34 (81%) | 5.86, 0.01 |
| Daily Surveys (Outside Visit) | 311 (63%) | 194 (55%) | 117 (80%) | 24.79, <0.01 |
| Weekly Surveys (Outside Visit) | 473 (74%) | 303 (67%) | 170 (90%) | 34.23, <0.01 |
| **Week 10 Visit Completed** | **53 (75%)** | **34 (68%)** | **19 (90%)** | **2.85, 0.09** |
| Interviews (During Visit) | 51 (72%) | 32 (64%) | 19 (90%) | 3.90, 0.04 |
| Dried Blood Spots (During Visit) | 44 (62%) | 30 (60%) | 14 (67%) | 0.07, 0.79 |
| Saliva Samples (During Visit) | 43 (61%) | 25 (50%) | 18 (86%) | 6.47, 0.01 |
| Saliva Samples (Outside Visit) | 81 (57%) | 45 (45%) | 36 (86%) | 18.38, <0.01 |
| Daily Surveys (Outside Visit) | 311 (63%) | 196 (56%) | 115 (78%) | 20.91, <0.01 |
| Weekly Surveys (Outside Visit) | 474 (74%) | 303 (67%) | 171 (90%) | 36.02, <0.01 |
| **Week 11 Visit Completed** | **53 (75%)** | **34 (68%)** | **19 (90%)** | **2.85, 0.09** |
| Interviews (During Visit) | 50 (70%) | 31 (62%) | 19 (90%) | 4.47, 0.03 |
| Dried Blood Spots (During Visit) | 47 (66%) | 31 (62%) | 16 (76%) | 0.77, 0.38 |
| Saliva Samples (During Visit) | 45 (63%) | 28 (56%) | 17 (81%) | 2.96, 0.08 |
| Saliva Samples (Outside Visit) | 90 (63%) | 55 (55%) | 34 (81%) | 7.44, <0.01 |
| Daily Surveys (Outside Visit) | 308 (62%) | 194 (55%) | 114 (78%) | 20.57, <0.01 |
| Weekly Surveys (Outside Visit) | 477 (75%) | 307 (68%) | 170 (90%) | 32.05, <0.01 |
| **Week 12 Visit Completed** | **53 (75%)** | **34 (68%)** | **19 (90%)** | **2.85, 0.09** |
| Interviews (During Visit) | 51 (72%) | 32 (64%) | 19 (90%) | 3.90, 0.05 |
| Dried Blood Spots (During Visit) | 45 (63%) | 30 (60%) | 15 (71%) | 0.41, 0.52 |
| Saliva Samples (During Visit) | 44 (62%) | 26 (52%) | 18 (86%) | 5.77, 0.02 |
| Saliva Samples (Outside Visit) | 89 (63%) | 53 (53%) | 36 (86%) | 12.17, <0.01 |
| Daily Surveys (Outside Visit) | 307 (62%) | 189 (54%) | 118 (80%) | 29.15, <0.01 |
| Weekly Surveys (Outside Visit) | 832 (73%) | 529 (66%) | 303 (90%) | 23.46, <0.01 |
| **Month 4 Visit Completed** | **52 (73%)** | **34 (68%)** | **18 (86%)** | **1.55, 0.21** |
| Interviews (During Visit) | 51 (72%) | 33 (66%) | 18 (86%) | 1.95, 0.16 |
| Dried Blood Spots (During Visit) | 46 (65%) | 29 (58%) | 17 (81%) | 2.48, 0.11 |
| Saliva Samples (During Visit) | 47 (66%) | 29 (58%) | 18 (86%) | 3.91, 0.05 |
| Saliva Samples (Outside Visit) | 90 (63%) | 56 (56%) | 34 (81%) | 6.89, <0.01 |
| Daily Surveys (Outside Visit) | 329 (66%) | 212 (61%) | 117 (80%) | 15.90, <0.01 |
| Weekly Surveys (Outside Visit) | 832 (73%) | 544 (68%) | 288 (86%) | 36.98, <0.01 |
| **Month 5 Visit Completed** | **52 (73%)** | **34 (68%)** | **18 (86%)** | **1.55, 0.21** |
| Interviews (During Visit) | 47 (66%) | 30 (60%) | 17 (81%) | 2.04, 0.15 |
| Dried Blood Spots (During Visit) | 44 (62%) | 29 (58%) | 15 (71%) | 0.63, 0.43 |
| Saliva Samples (During Visit) | 44 (62%) | 27 (54%) | 17 (81%) | 3.49, 0.06 |
| Saliva Samples (Outside Visit) | 90 (63%) | 56 (56%) | 34 (81%) | 6.89, <0.01 |
| Daily Surveys (Outside Visit) | 287 (58%) | 169 (48%) | 118 (80%) | 42.11, <0.01 |
| Weekly Surveys (Outside Visit) | 808 (71%) | 520 (65%) | 288 (86%) | 48.43, <0.01 |

* During the 7 days preceding birth.
