## Supplementary Document 1 for "Hormones and Infant Caregiving in Postpartum Opioid Use Disorder Recovery: Compliance and Satisfaction with a Prospective Cohort Study Protocol Designed to Identify Novel Support Targets"

**Supplementary Document 1. Lifetime Trauma and Resilience Interview**

**Interview Script, Questions, and Response Options (Completed at Baseline)**

I am going ask you some specific questions about things that may have happened when you were growing up and in your adult life. Some of these questions ask about difficult experiences you may have had in your life. We are interested in knowing more about these experiences so that we can begin to understand how parents are affected by and make sense of prior challenges, and how that process affects parenting. I am going to record our conversation today so that we can capture all the information you share with us. As always, we are very careful with your information, and we never use your name on our recordings. Is it alright with you if I record this part of the interview? Yes, no [If yes, start recording; If no, continue without recording]

These first questions ask about things that may have happened to you in the past 12 months. For each item, please answer no if it did not happen and yes if it did.

1. A close family member was very sick and had to go to the hospital. Yes, no, prefer not to answer
2. I got separated or divorced from my spouse or partner. Yes, no, prefer not to answer
3. I moved to a new address. Yes, no, prefer not to answer
4. I was homeless or had to sleep outside, in a car, or in a shelter. Yes, no, prefer not to answer
5. My spouse or partner lost their job. Yes, no, prefer not to answer
6. I lost my job even though I wanted to go on working. Yes, no, prefer not to answer
7. My spouse, partner, or I had a cut in work hours or pay. Yes, no, prefer not to answer
8. I was apart from my spouse or partner due to military deployment or extended work-related travel. Yes, no, prefer not to answer
9. I argued with my spouse or partner more than usual. Yes, no, prefer not to answer
10. My spouse or partner said they didn't want me to be pregnant. Yes, no, prefer not to answer
11. I had problems paying rent, mortgage, or other bills
12. My spouse, partner, or I went to jail. Yes, no, prefer not to answer
13. Someone very close to me had a problem with drinking or drugs. Yes, no, prefer not to answer
14. Someone very close to me died. Yes, no, prefer not to answer

Now, thinking about your whole life…

15. Were you ever exposed to a life-threatening natural disaster? Yes, no, prefer not to answer [If yes] How old were you? Age recorded
16. Were you involved in a serious accident? Yes, no, prefer not to answer [If yes] How old were you? Age recorded
17. Did you ever suffer a serious personal injury or illness? Yes, no, prefer not to answer [If yes] How old were you? Age recorded
18. Did you ever experience the death or serious illness of a parent or a primary caregiver? Yes, no, prefer not to answer [If yes] How old were you? Age recorded
19. Did you experience the divorce or separation of your parents? Yes, no, prefer not to answer [If yes] How old were you? Age recorded
20. Did you experience the death or serious injury of a sibling? Yes, no, prefer not to answer [If yes] How old were you? Age recorded
21. Did you ever experience the death or serious injury of a friend? Yes, no, prefer not to answer [If yes] How old were you? Age recorded
22. Did you ever witness violence towards others, including family members? Yes, no, prefer not to answer [If yes] How old were you? Age recorded
23. Did anyone in your family ever suffer from mental or psychiatric illness or have a "breakdown"? Yes, no, prefer not to answer [If yes] How old were you? Age recorded
24. Did your parents or primary caretaker have a problem with alcoholism or drug or drug abuse? Yes, no, prefer not to answer [If yes] How old were you? Age recorded
25. Did you ever see someone murdered? Yes, no, prefer not to answer [If yes] How old were you? Age recorded
26. Did a household member ever go to jail or prison? Yes, no, prefer not to answer [If yes] How old were you? Age recorded
27. Did your parents or adults in your home ever hit, punch, beat, or threaten to harm each other? Yes, no, prefer not to answer [If yes] How old were you? Age recorded

Next, I’d like to ask you about some other childhood experiences that may have an impact on you now, today. Sometimes people get spanked a lot, physically punished, or disciplined in a very strict way by their parents or caretakers when they are growing up. Before you were 18 …?

28. Were you ever slapped in the face with an open hand? <u>Yes*, no, prefer not to answer</u>
29. Were you ever burned with hot water, a cigarette or something else? <u>Yes*, no, prefer not to answer</u>
30. Were you ever punched or kicked? <u>Yes*, no, prefer not to answer</u>
31. Were you ever hit with an object that was thrown at you? <u>Yes*, no, prefer not to answer</u>
32. Were you ever pushed or shoved? <u>Yes*, no, prefer not to answer</u>
33. [Asked only if not already answered] Did a parent or other adult in the household: Push, grab, slap, or throw something at you? Or, ever hit you so hard that you had marks or were injured? <u>Yes*, no, prefer not to answer</u>

[* If yes, then the following were asked after each item endorsed.]

a. How old were you the first time something like this happened? <u>0-5, 6-10, 11-17</u>
b. How old were you the last time something like this happened? <u>0-5, 6-10, 11-17</u>
c. In general how often did this occur? <u>One time only, once a year, once a month, once a week, once a day</u>
d. Who is the person who did this the most? <u>Mother or mother figure, father or father figure, grandmother, grandfather, other female, other male</u>

While growing up people sometimes have sexual experiences that they didn’t want to have or that made them uncomfortable. Sometimes these experiences are with people they know and sometimes with strangers. Before the age of 18, did you ever experience any of the following things?

34. Were you ever touched in an intimate or private part of your body (e.g., breast, thighs, genitals) in a way that surprised you or made you feel uncomfortable? <u>Yes*, no, prefer not to answer</u>
35. Did you ever experience someone rubbing their genitals against you? <u>Yes*, no, prefer not to answer</u>
36. Were you ever forced or coerced to touch another person in an intimate or private part of their body? <u>Yes*, no, prefer not to answer</u>
37. Did anyone ever have genital sex with you against your will? <u>Yes*, no, prefer not to answer</u>
38. Were you ever forced or coerced to perform oral sex on someone against your will? <u>Yes*, no, prefer not to answer</u>
39. Were you ever forced or coerced to kiss someone in a sexual rather than an affectionate way? <u>Yes*, no, prefer not to answer</u>
40. [Asked only if not already answered] Did an adult or person at least 5 years older than you ever: touch or fondle you or have you touch their body in a sexual way? Or, attempt or actually have oral, anal, or vaginal intercourse with you? <u>Yes*, no, prefer not to answer</u>

[* If yes, then the following were asked after each item endorsed.]

a. How old were you the first time something like this happened? <u>0-5, 6-10, 11-17</u>
b. How old were you the last time something like this happened? <u>0-5, 6-10, 11-17</u>
c. In general how often did this occur? <u>One time only, once a year, once a month, once a week, once a day</u>
d. Who is the person who did this the most? <u>Mother or mother figure, father or father figure, grandmother, grandfather, other female, other male</u>

Sometimes while growing up people feel as if they can’t do anything right in their caregiver’s eyes. Their caregivers were always putting them down, yelling at them or telling them that they were no good. Before the age of 18 …?

41. Were you often put down or ridiculed? <u>Yes*, no, prefer not to answer</u>
42. Were you often ignored or made to feel that you didn’t count? <u>Yes*, no, prefer not to answer</u>
43. Were you often told you were no good? <u>Yes*, no, prefer not to answer</u>
44. Most of the time were you treated in a cold, uncaring way or made to feel like you were not loved? <u>Yes*, no, prefer not to answer</u>

45. Did your parents or caretakers often fail to understand you or your needs? Yes*, no, prefer not to answer
46. Did you often or very often feel that: no one in your family loved you or thought you were important or special? Or, your family didn't look out for each other, feel close to each other, or support each other? Yes*, no, prefer not to answer
47. Did a parent or other adult in the household often or very often act in a way that made you afraid that you might be physically hurt? Yes*, no, prefer not to answer

[* If yes, then the following were asked after each item endorsed].

a. How old were you the first time something like this happened? 0-5, 6-10, 11-17
b. How old were you the last time something like this happened? 0-5, 6-10, 11-17
c. In general how often did this occur? One time only, once a year, once a month, once a week, once a day
d. Who is the person who did this the most? Mother or mother figure, father or father figure, grandmother, grandfather, other female, other male

While growing up people may go through a period when they lack appropriate care (like not having enough to eat or drink, lacking shelter, being left alone when you were too young to care for yourself, or being left with a caregiver who was abusing drugs). Before age 18 …?

48. Were you ever left home alone when you felt that you were too young to care for yourself? Yes*, no, prefer not to answer
49. Was there a time when you felt there was not enough to eat or drink in your home? Yes*, no, prefer not to answer
50. Was there ever a time when you felt that your home did not provide adequate shelter (e.g., not sanitary, not safe, no working heat)? Yes*, no, prefer not to answer
51. Were you ever allowed to roam about outside unsupervised either at night or at another time when you were too young or you felt unsafe? Yes*, no, prefer not to answer
52. Was there ever a time when you were left with a caregiver who was abusing drugs or alcohol, or otherwise unable to care for you? Yes*, no, prefer not to answer
53. Was there ever a time when your caregiver(s) did not make sure you were regularly attending school or allowed you to be truant frequently, for extended periods of time? Yes*, no, prefer not to answer
54. [Ask only if not already answered] Did you often or very often feel that: you didn't have enough to eat, had to wear dirty clothes, and had no one to protect you? Or your parents were too drunk or high to take care of you or take you to the doctor if you needed it? Yes*, no, prefer not to answer

[* If yes, then the following were asked after each item endorsed.]

a. How old were you the first time something like this happened? 0-5, 6-10, 11-17
b. How old were you the last time something like this happened? 0-5, 6-10, 11-17
c. In general how often did this occur? One time only, once a year, once a month, once a week, once a day
d. Who is the person who did this the most? Mother or mother figure, father or father figure, grandmother, grandfather, other female, other male

As you think about the events we have been talking about related to negative physical, emotional, or sexual experiences in your childhood…

55. Do you believe that these events have an emotional effect on you today? Yes**, no, prefer not to answer
56. Do you believe that these events affect your current functioning at work or school? Yes**, no, prefer not to answer
57. Do you believe these events affect your current social or family relationships? Yes**, no, prefer not to answer
58. Do you believe these events affect, or will affect, how you function as a caregiver? Yes**, no, prefer not to answer

[** If yes for questions 53-60, then the following was asked after each endorsed item.] What kind of effect? Extremely negative, moderately negative, slightly negative, slightly positive, moderately positive, extremely positive

We've been talking about some difficult things just now and I want to thank you for your willingness to share your experiences. Next, I'm going to ask about your feelings and beliefs about important relationships in your life while you were growing up.

59. I believe that my mother loved me when I was little. Definitely not true, probably not true, not sure, probably true, definitely not true, prefer not to answer
60. I believe that my father loved me when I was little. Definitely not true, probably not true, not sure, probably true, definitely not true, prefer not to answer
61. When I was little, other people helped my parents take care of me and they seemed to love me. Definitely not true, probably not true, not sure, probably true, definitely not true, prefer not to answer
62. I've heard that when I was an infant, someone in my family enjoyed playing with me and I enjoyed it too. Definitely not true, probably not true, not sure, probably true, definitely not true, prefer not to answer
63. When I was a child, there were relatives in my family who helped me feel better when I was sad or worried. Definitely not true, probably not true, not sure, probably true, definitely not true, prefer not to answer
64. When I was a child, neighbors or my friends' parents seemed to like me. Definitely not true, probably not true, not sure, probably true, definitely not true, prefer not to answer
65. When I was a child, teachers, coaches, youth leaders, or ministers were there to help me. Definitely not true, probably not true, not sure, probably true, definitely not true, prefer not to answer
66. Someone in my family cared about how I was doing in school. Definitely not true, probably not true, not sure, probably true, definitely not true, prefer not to answer
67. My family, friends, and neighbors talked about making our lives better. Definitely not true, probably not true, not sure, probably true, definitely not true, prefer not to answer
68. We had rules in our house and were expected to keep them. Definitely not true, probably not true, not sure, probably true, definitely not true, prefer not to answer
69. When I felt bad, I could almost always find someone I trusted to talk to. Definitely not true, probably not true, not sure, probably true, definitely not true, prefer not to answer
70. As a youth, people noticed that I was capable and could get things done. Definitely not true, probably not true, not sure, probably true, definitely not true, prefer not to answer
71. I was independent and a go-getter. Definitely not true, probably not true, not sure, probably true, definitely not true, prefer not to answer
72. I believe that life is what you make it. Definitely not true, probably not true, not sure, probably true, definitely not true, prefer not to answer
73. There are people I can count on now in my life. Definitely not true, probably not true, not sure, probably true, definitely not true, prefer not to answer

Thank you again for your willingness to share your experiences. Now I want to ask you a few questions your hopes and dreams for your baby.

74. If you had three wishes for your baby's future, what would they be? [open field]
75. What do you think are the three most important things that you can offer your baby? [open field]
76. Is there anything else you'd like to share with us? [open field]

---

Specific Items Link to Validated and Investigator-Created Measures

Stressful life events (Newton et al[32]): Items 1-14

Modified Early Trauma Inventory (Bremner et al [34,35]): General Traumas: Items 15-27, Physical Punishment: Items 28-33, Sexual Events: Items 34-40, Emotional Abuse: Items 41-47, Neglect: Items 48-54, Impact: Items 55-58

Adverse Childhood Experiences (Felitti et al[33]): Coded from Early Trauma Inventory Items 18, 19, 22-24, 26-54

Resilience Scale (Wagnild et al[36]): Items 59-73

Reflection Questions (investigator-created): Items 74-76
