## Supplementary Document 2 for "Hormones and Infant Caregiving in Postpartum Opioid Use Disorder Recovery: Compliance and Satisfaction with a Prospective Cohort Study Protocol Designed to Identify Novel Support Targets"

**Supplementary Document 2. Investigator-Created Birth and Breastfeeding Interview**

**Birth Interview Script, Questions, and Response Options (Completed at Postpartum Week 1)**

I'd like to ask you some questions ask about your pregnancy and your child's birth. I am going to record our conversation today so that we can capture all the information you share with us. As always, we are very careful with your information, and we never use your name on our recordings. Is it alright with you if I record this part of the interview? Yes/No [If yes, start recording; If no, continue without recording]

First, I'd like you to think back to the time you discovered you were pregnant…

1. How far along in the pregnancy were you when you first found out you were pregnant? [open response]
2. Was this a planned pregnancy? [open response]
3. Tell me the first thing you felt and thought when you found out you were pregnant? [open response]
4. How did discovering you were pregnant change things for you? [open response]
5. Overall, how was your pregnancy? [open response]
   a. [Prompt if needed] Who or what was a challenge for you during the pregnancy? [open response]
   b. [Prompt if needed] Who or what was a source of support for you? [open response]
6. How was your interaction with the healthcare system during your pregnancy? [open response]

Now, let's talk about your baby's birth and what it was like for you after you had your baby. We are particularly interested in your birth experience and any medications or interventions you or (infant's name) had during or right after the birth. This is because your experiences during birth can influence your postpartum hormones and the feelings you have now.

7. When was your baby born? [open response]
8. Tell me the first thing you felt and thought when (infant's name) was first delivered? [open response]
9. Where did you give birth? [open response]
10. Did you have any support people present with you for the birth? Who? [open response]
11. Was your delivery vaginal or by cesarean section? Vaginal, Cesarean Section, Refused
    a. [If cesarean section] Was this a planned or emergency surgery? [Open response]
12. Did you go into labor spontaneously or were you induced? Spontaneously, Induced, Unsure
    a. [If spontaneous] Did you receive Pitocin or other medication to augment your labor at any point? [open response]
    b. [If induced] What method was used (e.g., Pitocin; other medication; non-pharmacological method such as balloon expansion or membrane sweep)? [open response]
13. Did you receive medication at any point during labor or after the birth? [open response]
    a. [Prompt if needed] Do you know if you received Pitocin or another medication to assist with birthing the placenta/afterbirth? [open response]
    b. [Prompt if needed] Were you prescribed any new medications or had any changes to your medications (e.g., pain medication for cesarean recovery; adding or resuming medication after the birth)? [open response]
14. Did you or (infant's name) experience any complications during labor or birth (e.g., hypertension, forceps, hypoxia)? [open response]
15. Was there ever a point that you felt concerned for yourself or your baby? [open response]
16. How much did (infant's name) weigh at birth? [open response]
17. Overall, how was it for you while you were in labor? [open response]
    a. [Prompt if needed] What was helpful or not helpful? [open response]
    b. [Prompt if needed] How was the hospital staff? [open response]
18. Overall, how was your experience of the birth? [open response]

Thinking about your experiences after the birth…

19. How long did you stay in the hospital or birthing center? [open response]
20. Did you spend the first hours/days after birth with your baby (e.g., rooming in)? [open response]
21. Were you separated from your baby at any point after birth? [open response]
22. Were you and (infant's name) discharged at the same time? [open response]
23. Did (infant's name) require specialized care or admission to the NICU? Did they receive any medication? [open response]
24. Did you initiate or attempt to initiate breastfeeding? Yes, no
    a. [If yes] When did you first breastfeed? [open response]

b. Did you experience any difficulties with breastfeeding? [open response]
c. Did you receive breastfeeding support from a doula, lactation consultant, or other professional? [open response]

Now I'm going to ask you about your experiences since coming home from the hospital...

25. Are you currently breastfeeding and/or pumping milk for your baby? Yes, no, still initiating breastfeeding, other.
    a. [If participant endorses any breastfeeding, pumping, or intention to breastfeed] What are your top three reasons for breastfeeding your baby or pumping milk for your baby? [open response]
    b. Was there anyone who encouraged or supported you to breastfeed? [open response]
    c. Did anyone want you to stop breastfeeding? [open response]
    d. How enjoyable was/is your experience of breastfeeding your baby? [answered on a 100-point VAS scale where 0 is "Not at all" and 100 is "Very"]
    e. How challenging was/is your experience of breastfeeding your baby? [answered on a 100-point VAS scale where 0 is "Not at all" and 100 is "Very"]
    f. How likely is it that you would breastfeed again if you had another child? [answered on a 100-point VAS scale where 0 is "Not at all" and 100 is "Very"]
26. How have things been in these first days since you and (infant's name) came home? [open response]
    a. How have you been feeling and adjusting? [open response]
    b. What kind of supports and resources do you have? [open response]
    c. What has been challenging for you? [open response]
    d. What has been a source of joy? [open response]

### Breastfeeding Interview Script, Questions, and Response Options (Completed at Postpartum Week 1 and Postpartum Month 5)

[Continued from above for Postpartum Week 1 without reading the following script; Postpartum Month 5 reads the following to start.] I'd like to ask you some questions ask about your pregnancy and your child's birth. I am going to record our conversation today so that we can capture all the information you share with us. As always, we are very careful with your information, and we never use your name on our recordings. Is it alright with you if I record this part of the interview? Yes/No [If yes, start recording; If no, continue without recording]

1. Are you breastfeeding and/or pumping milk for your baby? Yes, no, still initiating breastfeeding, other
    a. [If yes] What are your top three reasons for breastfeeding your baby or pumping milk for your baby? [open response]
    b. [At postpartum week 1 only, if no] Did you initiate breastfeeding or attempt to breastfeed at any time since your baby was born? [open response]
        i. [If yes] Did you breastfeed as long as you wanted to? [open response]
        ii. [if yes] What were the top three reasons for your decision to stop breastfeeding your baby? [open response]
2. Was there anyone who encouraged or supported you to breastfeed? [open response]
    a. [Probe for significant relationships if needed including: The baby's father; participant's romantic partner; participant's mother; participant's mother-in-law; participant's grandparents or extended family; doctor or other healthcare professional; supervisor or employer]
3. Did anyone want you to stop breastfeeding? [open response]
    a. [Probe for significant relationships if needed including: The baby's father; participant's romantic partner; participant's mother; participant's mother-in-law; participant's grandparents or extended family; doctor or other healthcare professional; supervisor or employer]
4. How enjoyable was/is experience of breastfeeding your baby? [answered on a 100-point VAS scale where 0 is "Not at all enjoyable" and 100 is "Very enjoyable"]
5. How challenging was/is experience of breastfeeding your baby? [answered on a 100-point VAS scale where 0 is "Not at all challenging" and 100 is "Very challenging"]
6. How likely is it that you would breastfeed again if you had another child? [answered on a 100-point VAS scale where 0 is "Not at all likely" and 100 is "Very likely"]
