## Supplementary Document 3 for "Hormones and Infant Caregiving in Postpartum Opioid Use Disorder Recovery: Compliance and Satisfaction with a Prospective Cohort Study Protocol Designed to Identify Novel Support Targets"

**Supplementary Document 3. Investigator-Created and Modified Versions of Validated Instruments**

Sociodemographics

[Completed once at Baseline. Note: All responses will include a “prefer not to answer” response.]

1. What is your date of birth? _______
2. What is your current housing status? Check all that apply
    a. Rent/own
    b. Living with a husband or partner
    c. Living with a family member
    d. Living with a friend
    e. At risk for losing housing
    f. Living in a shelter
    g. Living in transitional housing
    h. Homeless
    i. Other, describe: ___________
3. What is your present marital status?
    a. Never married
    b. Partnered but not married
    c. Married for the first time
    d. Separated
    e. Divorced
    f. Remarried
    g. Widowed
    h. Other, describe: ___________
4. When you got pregnant, what relationship did you have with your new baby’s father?
    a. He was my husband (legally married)
    b. He was my partner (not legally married)
    c. He was my boyfriend
    d. He was my friend
    e. We had no relationship
    f. Other, describe: ___________
5. Now, what relationship do you have with your new baby’s father?
    a. He is my husband (legally married)
    b. He is my partner (not legally married)
    c. He is my boyfriend
    d. He is my friend
    e. We do not have a relationship
    f. Other, describe: ___________
6. How many children are currently living in your household? ____
7. How many adults are currently living in your household? ____

Medical History
[Completed once at Baseline. Note: All responses will include a “prefer not to answer” response.]

1. When was the first day of your last menstrual period? ______
2. When are you due to have your baby? __________
3. Including this one, how many times have you been pregnant? ___
4. How many times have you given birth (at least gestational week 24)? ____
5. For this pregnancy, how many weeks or months pregnant were you when you had you first visit for prenatal care? ____ weeks or ____ months
6. Are you planning to breastfeed once your baby is born?
    a. Yes, exclusively
    b. Yes, but alternating with formula
    c. No, not at all
    d. Undecided
7. During this pregnancy or your past pregnancies, have you been told you have any of the following: (Response options for each are yes, no, don’t know, prefer not to answer; for any ‘yes’ response text boxes will appear for ‘when? ___’ and ‘describe: ____’)
    a. Gestational diabetes
    b. High blood pressure
    c. Fetal growth restriction
    d. Multiple fetuses
    e. Congenital anomaly
    f. Any other problems
8. Have you ever experienced or been diagnosed with any of the following: (Response options for each are yes, no, don’t know, prefer not to answer; for any ‘yes’ response text boxes will appear for ‘when? ___’ and ‘describe: ____’)
    a. Ovarian hypofunction
    b. Polycystic ovarian syndrome
    c. Premenstrual dysphoric disorder
    d. Endometriosis
    e. Rapid weight loss
    f. Low body fat
    g. Klinefelter syndrome
    h. Turner syndrome
    i. Adrenal disorders
    j. Alcoholism
    k. Hemochromatosis
    l. Sickle cell disease
    m. Depression
    n. Pain that occurred every day and lasted for 3 months or longer
9. Are you currently using any of the following: (Response options for each are yes, no, don’t know, prefer not to answer; for any ‘yes’ response text boxes will appear for ‘when? ___’ and ‘describe: ____’)
    a. Hormonal contraceptives
    b. Hormones
    c. St. John’s Wort
    d. Phenobarbital
    e. Carbamazepine
    f. Rifampicin
    g. Erythromycin or clarithromycin
    h. Ketoconazole or itraconazole
    i. Ritonavir
    j. Grapefruit juice
    k. Clomiphene
    l. Naloxone
    m. Digoxin

10. After you have your baby do you intend to use any of the following: (Response options for each are yes, no, don't know, prefer not to answer; for any 'yes' or 'don't know' response text boxes will appear for 'describe: ____')
    a. Hormonal contraceptives
    b. Hormones
    c. St. John's Wort
    d. Phenobarbital
    e. Carbamazepine
    f. Rifampicin
    g. Erythromycin or clarithromycin
    h. Ketoconazole or itraconazole
    i. Ritonavir
    j. Grapefruit juice
    k. Clomiphene
    l. Naloxone
    m. Digoxin
11. What medications are you currently taking? Be sure to include any prescription and nonprescription medications, as well as any vitamins or supplements. (Collect name of medication, reason for use, dose, when started)
12. Do you have any major health issues or concerns? ______________________________

Substance Use History
[Completed once on REDCap at Baseline and #20-25 will be administered weekly beginning at postpartum Week 1 and again at Month 4 and Month 5. Note: All responses will include a "prefer not to answer" response.]

1. Have you ever used any of the following drugs for any reason? For each item, check No if you did not use it or Yes if you did.
    a. Caffeine (including coffee, teas, soda, and other beverages that contain caffeine)
    b. Non-opiate pain relievers including over-the-counter medication such as aspirin, Tylenol®, Advil®, or Aleve® or prescription strength ibuprofen or acetaminophen
    c. Opiate-containing prescription pain relievers such as hydrocodone (Vicodin ®), oxycodone (Percocet ®), or codeine
    d. Prescription antidepressants or selective serotonin reuptake inhibitors (SSRIs) such as Prozac, Zoloft, or Lexapro, or prescription mood stabilizers such as lithium or Lamictal
    e. Adderall ®, Ritalin ®, or another stimulant
    f. Cigarettes, e-cigarettes, or other forms of tobacco or nicotine
    g. Alcohol, including beer, wine, or hard liquor
    h. Marijuana or hash
    i. Synthetic marijuana (K2, spice)
    j. Methadone, naloxone, Subutex, or Suboxone ®
    k. Heroin (smack, junk, Black Tar, Chiva)
    l. Amphetamines (uppers, speed, crystal meth, crank, ice, agua)
    m. Cocaine (crack, rick, coke, blow, snow, nieve)
    n. Tranquilizers (downers, ludes)
    o. Hallucinogens (LSD/acid, PCP/angel dust, Ecstasy, Molly, mushrooms, bath salts)
    p. Sniffing gasoline, glue, aerosol spray cans, or paint to get high (huffing)
    q. Other
        i. If yes, describe: __________
2. [If 1c selected] How old were you the first time you used prescription pain relievers? ___ years
3. [If 1c selected] When was the last time you used prescription pain relievers? ___ days, months, or years ago
4. [If 1c selected] What type of prescription pain reliever did you last use? _______________________
5. [If 1c selected] How much were you using the last time you used? ___________
6. [If 1k selected] How old were you the first time you used heroin (smack, junk, Black Tar, Chiva)? ___ years
7. [If 1k selected] When was the last time you used heroin (smack, junk, Black Tar, Chiva)? ___ days, months, or years
8. [If 1k selected] How were you using heroin (smack, junk, Black Tar, Chiva) the last use? ___________
9. [If 1k selected] How much were you using the last time you used? ___________
10. During the three months before you got pregnant, did you take or use any of the following drugs for any reason? For each item, check No if you did not use it or Yes if you did.
    a. Caffeine (including coffee, teas, soda, and other beverages that contain caffeine)
    b. Non-opiate pain relievers including over-the-counter medication such as aspirin, Tylenol®, Advil®, or Aleve® or prescription strength ibuprofen or acetaminophen
    c. Opiate-containing prescription pain relievers such as hydrocodone (Vicodin ®), oxycodone (Percocet ®), or codeine
    d. Prescription antidepressants or selective serotonin reuptake inhibitors (SSRIs) such as Prozac, Zoloft, or Lexapro, or prescription mood stabilizers such as lithium or Lamictal
    e. Adderall ®, Ritalin ®, or another stimulant
    f. Cigarettes, e-cigarettes, or other forms of tobacco or nicotine
        i. If yes, how much?
    g. Alcohol, including beer, wine, or hard liquor
    h. Marijuana or hash
    i. Synthetic marijuana (K2, spice)
    j. Methadone, naloxone, Subutex, or Suboxone ®
    k. Heroin (smack, junk, Black Tar, Chiva)
    l. Amphetamines (uppers, speed, crystal meth, crank, ice, agua)
    m. Cocaine (crack, rick, coke, blow, snow, nieve)

    n. Tranquilizers (downers, ludes)
    o. Hallucinogens (LSD/acid, PCP/angel dust, Ecstasy, Molly, mushrooms, bath salts)
    p. Sniffing gasoline, glue, aerosol spray cans, or paint to get high (huffing)
    q. Other; If yes, describe: __________

11. During the <u>last three months of your most recent pregnancy</u>, did you take or use any of the following drugs for any reason? For each item, check No if you did not use it or Yes if you did.
    a. Caffeine (including coffee, teas, soda, and other beverages that contain caffeine)
    b. Non-opiate pain relievers including over-the-counter medication such as aspirin, Tylenol®, Advil®, or Aleve® or prescription strength ibuprofen or acetaminophen
    c. Opiate-containing prescription pain relievers such as hydrocodone (Vicodin ®), oxycodone (Percocet ®), or codeine
    d. Prescription antidepressants or selective serotonin reuptake inhibitors (SSRIs) such as Prozac, Zoloft, or Lexapro, or prescription mood stabilizers such as lithium or Lamictal
    e. Adderall®, Ritalin® or another stimulant
    f. Cigarettes, e-cigarettes, or other forms of tobacco or nicotine
        i. If yes, how much?
    g. Alcohol, including beer, wine, or hard liquor
    h. Marijuana or hash
    i. Synthetic marijuana (K2, Spice)
    j. Methadone, naloxone, Subutex, or Suboxone®
    k. Heroin (smack, junk, Black Tar, Chiva)
    l. Amphetamines (uppers, speed, crystal meth, crank, ice, agua)
    m. Cocaine (crack, rock, coke, blow, snow, nieve)
    n. Tranquilizers (downers, ludes)
    o. Hallucinogens (LSD/acid, PCP/angel dust, Ecstasy, Molly, mushrooms, bath salts)
    p. Sniffing gasoline, glue, aerosol spray cans, or paint to get high (huffing)
    q. Other; If yes, describe: __________

12. <u>Now</u>, do you take or use any of the following drugs for any reason? For each item, check No if you did not use it or Yes if you did.
    a. Caffeine (including coffee, teas, soda, and other beverages that contain caffeine)
    b. Non-opiate pain relievers including over-the-counter medication such as aspirin, Tylenol®, Advil®, or Aleve® or prescription strength ibuprofen or acetaminophen
    c. Opiate-containing prescription pain relievers such as hydrocodone (Vicodin ®), oxycodone (Percocet ®), or codeine
    d. Prescription antidepressants or selective serotonin reuptake inhibitors (SSRIs) such as Prozac, Zoloft, or Lexapro, or prescription mood stabilizers such as lithium or Lamictal
    e. Adderall®, Ritalin® or another stimulant
    f. Cigarettes, e-cigarettes, or other forms of tobacco or nicotine
        i. If yes, how much?
    g. Alcohol, including beer, wine, or hard liquor
    h. Marijuana or hash
    i. Synthetic marijuana (K2, Spice)
    j. Methadone, naloxone, Subutex, or Suboxone®
    k. Heroin (smack, junk, Black Tar, Chiva)
    l. Amphetamines (uppers, speed, crystal meth, crank, ice, agua)
    m. Cocaine (crack, rock, coke, blow, snow, nieve)
    n. Tranquilizers (downers, ludes)
    o. Hallucinogens (LSD/acid, PCP/angel dust, Ecstasy, Molly, mushrooms, bath salts)
    p. Sniffing gasoline, glue, aerosol spray cans, or paint to get high (huffing)
    q. Other; If yes, describe: __________

13. Overall, which is your drug of choice? Select one.
    a. Caffeine (including coffee, teas, soda, and other beverages that contain caffeine)
    b. Opiate-containing prescription pain relievers such as hydrocodone (Vicodin®), oxycodone (Percocet®), or codeine
    c. Adderall®, Ritalin® or another stimulant
    d. Cigarettes, e-cigarettes, or other forms of tobacco or nicotine

        i. If yes, how much?
    e. Alcohol, including beer, wine, or hard liquor
    f. Marijuana or hash
    g. Synthetic marijuana (K2, Spice)
    h. Methadone, naloxone, Subutex, or Suboxone®
    i. Heroin (smack, junk, Black Tar, Chiva)
    j. Amphetamines (uppers, speed, crystal meth, crank, ice, agua)
    k. Cocaine (crack, rock, coke, blow, snow, nieve)
    l. Tranquilizers (downers, ludes)
    m. Hallucinogens (LSD/acid, PCP/angel dust, Ecstasy, Molly, mushrooms, bath salts)
    n. Sniffing gasoline, glue, aerosol spray cans, or paint to get high (huffing)
    o. Other; If yes, describe: __________

14. [If 1c, 1j, and/or 1k selected] Which of the following have ever been true for you? Check ALL that apply
    a. Taking an opioid in larger amounts than prescribed or intended
    b. Taking an opioid for a longer time than prescribed or intended
    c. Wanting to cut down or quit but not being able to do so
    d. Spending a lot of time obtaining the opioid
    e. Having a craving or strong desire to use opioids
    f. Repeatedly unable to carry out major obligations (such as at work, at school, or at home) due to opioid use
    g. Continued use of opioids even though it caused or worsened social or relationship problems
    h. Stopping or reducing important social or work activities due to opioid use
    i. Recurrent use of opioids in physically hazardous situation (such as driving or at work)
    j. Continuing to use opioids even though you experienced physical or psychosocial difficulties from using opioids
15. [For OUD+ only.] What type of treatment have you ever tried for your opioid use disorder? Check ALL that apply
    a. Inpatient
        a. If selected: Was this (check all that apply): legally mandated, encouraged by friends and family, pursued on your own
    b. Intensive Outpatient (IOP)
        a. If selected: Was this (check all that apply): legally mandated, encouraged by friends and family, pursued on your own
    c. Outpatient
        a. If selected: Was this (check all that apply): legally mandated, encouraged by friends and family, pursued on your own
16. [For OUD+ only.] What type of treatment are you in now for your opioid use disorder? Check ALL that apply
    a. Inpatient
    b. Intensive Outpatient (IOP)
    c. Outpatient
    d. Counseling or Support
        i. Individual behavioral counseling
        ii. Group behavioral counseling
        iii. Peer support
        iv. 12-step or similar recovery program
        v. Telephone-based counseling or support
        vi. Online-based counseling or support
        vii. Text messaging counseling or support
    e. Medications
        viii. Methadone
        ix. Buprenorphine (Subutex)
        x. Buprenorphine/Naloxone (Suboxone)
        xi. Naltrexone, oral
        xii. Naltrexone, injectable
        xiii. Other, describe: ____________

    f. Other, describe: ____________
    g. None
17. [For OUD+ only.] How long have you been in recovery? Recovery can be defined as a process of change towards health and wellness. In this case, we are interested in when you started the process of change from regularly using opioids to stopping (or when you switched to a maintenance medication such as methadone or buprenorphine).
    a. Several years or more
    b. Within the past year but before I become pregnant
    c. Since becoming pregnant:
        i. during my first trimester
        ii. during my second trimester
        iii. during my third trimester
    d. Something else, describe: ____________
18. [For OUD+ only.] At this time, which of the following best describes your personal goal with regard to recovery after you delivery your baby?
    a. To remain in recovery and active in your treatment program
    b. To remain in recovery and discontinue your treatment program
    c. To use your drug of choice, but only once in awhile
    d. To resume use of your drug of choice
    e. You are not sure what your goal is right now
19. [For OUD+ only How likely are you to not use your drug of choice in the first nine months after baby is born?
    a. Extremely likely
    b. Very likely
    c. Somewhat likely
    d. Not very likely
    e. Not at all likely
20. [Timeline Followback procedure to be administered interview-style weekly beginning at postpartum Week 1 and again at Month 4 and Month 5] In the past week [or past month], have you taken or used any of the following drugs for any reason? For each item, check No if you did not use it or Yes if you did.
    a. Caffeine (including coffee, teas, soda, and other beverages that contain caffeine)
    b. Non-opiate pain relievers including over-the-counter medication such as aspirin, Tylenol®, Advil®, or Aleve® or prescription strength ibuprofen or acetaminophen
    c. Opiate-containing prescription pain relievers such as hydrocodone (Vicodin ®), oxycodone (Percocet ®), or codeine
    d. Prescription antidepressants or selective serotonin reuptake inhibitors (SSRIs) such as Prozac, Zoloft, or Lexapro, or prescription mood stabilizers such as lithium or Lamictal
    e. Adderall®, Ritalin® or another stimulant
    f. Cigarettes, e-cigarettes, or other forms of tobacco or nicotine
    g. Alcohol, including beer, wine, or hard liquor
    h. Marijuana or hash
    i. Synthetic marijuana (K2, Spice)
    j. Methadone, naloxone, Subutex, or Suboxone®
    k. Heroin (smack, junk, Black Tar, Chiva)
    l. Amphetamines (uppers, speed, crystal meth, crank, ice, agua)
    m. Cocaine (crack, rock, coke, blow, snow, nieve)
    n. Tranquilizers (downers, ludes)
    o. Hallucinogens (LSD/acid, PCP/angel dust, Ecstasy, Molly, mushrooms, bath salts)
    p. Sniffing gasoline, glue, aerosol spray cans, or paint to get high (huffing)
    q. Other; If yes, describe: __________

    [When any item is selected “yes” on #20 (with the exception of caffeine, non-opiate pain relievers, SSRIs), the following items are asked for each substance endorsed for each of the previous 7 days.]
21. Did you use/take ____________ on _______ [each date starting with “today” and going back for 7 [or 30] consecutive days; if asking about cigarettes, e-cigarettes, or other forms of tobacco or nicotine, follow-up question will include “What type of tobacco product did you use]? [Yes/No]

22. [Asked for each substance and day endorsed as “yes” on #21] How much of _____________ did you use/take on [date]?  Dose/amount used: ______ Frequency used: _____
23. [Asked for each substance endorsed as “yes” on #21]  Was ____________ prescribed by a healthcare provider? [Yes/No] Dose prescribed: ______ Frequency prescribed: ______
24. [Asked for each item selected as “yes” on #20] Do you believe you overused or misused __________ at any time during the past week
25. [If “yes” on #24] Why do you believe you overused/misused ___________? [Allows for open-ended response; probe for antecedents including stressors, significant events, relationship changes, etc.]

Daily Assessment

(Completed daily beginning the day after baseline until postpartum week 12, and then daily for seven days again at postpartum months 4, and 5. Additionally, #1-3 will be completed at baseline. All items will include a "prefer not to answer" option)

1. [prior to child birth only] Have you had your baby? [Yes/No]
2. Thinking about the past 24 hours, did you crave any of the following today? [Specific items listed are those endorsed in Substance Use History #10-13; respond with 100-point VAS scale where 0 is not at all and 100 is severe craving or prefer not to answer]
    a. Caffeine (including coffee, teas, soda, and other beverages that contain caffeine)
    b. Opiate-containing prescription pain relievers such as hydrocodone (Vicodin ®), oxycodone (Percocet ®), or codeine
    c. Adderall®, Ritalin® or another stimulant
    d. Cigarettes, e-cigarettes, or other forms of tobacco or nicotine
    e. Alcohol, including beer, wine, or hard liquor
    f. Marijuana or hash
    g. CBD oil
    h. Synthetic marijuana (K2, Spice)
    i. Methadone, naloxone, Subutex, or Suboxone®
    j. Heroin (smack, junk, Black Tar, Chiva)
    k. Amphetamines (uppers, speed, crystal meth, crank, ice, agua)
    l. Cocaine (crack, rock, coke, blow, snow, nieve)
    m. Tranquilizers (downers, ludes)
    n. Hallucinogens (LSD/acid, PCP/angel dust, Ecstasy, Molly, mushrooms, bath salts)
    o. Sniffing gasoline, glue, aerosol spray cans, or paint to get high (huffing)
    p. [presented for everyone] Any [other] substance
        i. If selected, then: which did you crave? [full list from above is presented]
3. Did you use any of the following today? [Specific items listed are those endorsed in Substance Use History #10-13. If no endorsements, participants are asked about any use [item r]; response options include: no, yes, prefer not to answer]
    a. Caffeine (including coffee, teas, soda, and other beverages that contain caffeine)
    b. Non-opiate pain relievers including over-the-counter medication such as aspirin, Tylenol®, Advil®, or Aleve® or prescription strength ibuprofen or acetaminophen
    c. Opiate-containing prescription pain relievers such as hydrocodone (Vicodin ®), oxycodone (Percocet ®), or codeine
    d. Prescription antidepressants or selective serotonin reuptake inhibitors (SSRIs) such as Prozac, Zoloft, or Lexapro
    e. Adderall®, Ritalin® or another stimulant
    f. Cigarettes, e-cigarettes, or other forms of tobacco or nicotine
        i. If yes, how much? _______
    g. Alcohol, including beer, wine, or hard liquor
    h. Marijuana or hash
    i. CBD oil
    j. Synthetic marijuana (K2, Spice)
    k. Methadone, naloxone, subutex, or Suboxone®
    l. Heroin (smack, junk, Black Tar, Chiva)
    m. Amphetamines (uppers, speed, crystal meth, crank, ice, agua)
    n. Cocaine (crack, rock, coke, blow, snow, nieve)
    o. Tranquilizers (downers, ludes)
    p. Hallucinogens (LSD/acid, PCP/angel dust, Ecstasy, Molly, mushrooms, bath salts)
    q. Sniffing gasoline, glue, aerosol spray cans, or paint to get high (huffing)
    r. [presented for everyone] Any [other] substance
        i. If selected, then: which did you use? [full list from above is presented]
4. [Presented to those who endorsed any level of craving for any substance] Overall, how well do you think you coped with your cravings today? [respond with 100-point VAS scale where 0 is not at all and 100 is very well or prefer not to answer]

5. [Presented to those who endorsed any level of craving for any substance and did not use; randomly launched for a maximum of one time per week] We are interested in how you cope with cravings. Do you want to share? Please list a strategy you used today to avoid using even though you had cravings.
6. [Presented until postpartum week 4 only] We are interested in whether you are experiencing pain on a regular basis. Please rate your pain at its <u>worst</u> in the last 24 hours. [respond with 100-point VAS scale where 0 is none at all and 100 is pain as bad as you can imagine or prefer not to answer]
7. [Presented until postpartum week 4 only] Please rate your pain <u>on average</u> in the last 24 hours. [respond with 100-point VAS scale where 0 is none at all and 100 is pain as bad as you can imagine or prefer not to answer]
8. [postpartum only] Were you with your baby today? [yes/no]
9. [postpartum only; if yes to #8 only] Today, how much time did you spend… (answered on a 100-point VAS scale where 0 is "A lot less than usual" and 100 is "A lot more than usual")
    a. In skin-to-skin contact with your baby (including holding unclothed baby on your skin, breastfeeding, etc.)?
    b. In other physical contact with your baby (including changing your baby, holding your baby, feeding your baby, babywearing, etc.)?
    c. Caring for your baby but not in physical contact (including baby playing, tummy time, baby in a swing or car seat, baby sleeping in crib, etc.)?
    d. With someone else while they cared for your baby (e.g., dad changing baby, grandma feeding baby)?
    e. Away from your baby while your baby was being cared for by someone else (e.g., another family member, child care center, nanny, etc. while you were at work or school)?
10. [postpartum only; if yes to #8 only] (Daily question for first four weeks) How did you feed your baby today? (OR weekly question thereafter) How did you feed your baby during the past week?
    a. Only with my breastmilk
    b. Mostly with my breastmilk and some formula
    c. About equally with my breastmilk and formula
    d. Mostly with formula and some with my breastmilk
    e. Only with formula
    f. Other (Describe: _____________)
11. [postpartum only; if yes to #8 only] For the times when you baby is receiving your breastmilk, how are you giving your baby your breastmilk:
    a. Breastfeeding only
    b. A combination of breastfeeding and pumping
    c. Pumping only

[postpartum only; if yes to #8 only] Thinking about all the things that are involved in taking care of your baby…

12. [postpartum only; if yes to #8 only] How challenging did you find parenting today? (answered on a 100-point VAS scale where 0 is "not at all" and 100 is "extremely")
13. [postpartum only; if yes to #8 only] How rewarding did you find parenting today? (answered on a 100-point VAS scale where 0 is "not at all" and 100 is "extremely")

Thinking about your connections to people…

14. How connected to your baby did you feel today? (answered on a 100-point VAS scale where 0 is "not at all" and 100 is "extremely")
15. How connected did you feel with others today? (answered on a 100-point VAS scale where 0 is "not at all" and 100 is "extremely")
16. [Starting at postpartum week 4] Finally, did you have any vaginal bleeding today?
    a. No, not at all
    b. Yes, light spotting
    c. Yes, about the same as a menstrual period
    d. Yes, heavier than a menstrual period
    e. Other (Describe: ______________)

Modified Pregnancy and Postpartum Pain Inventory
(Completed weekly beginning at baseline and continuing until postpartum week 4, and then again at postpartum months 4 and 5. All items will include a "prefer not to answer" option.)

1. It's common to have pain during pregnancy and after having a baby. Have you experienced pain other than everyday pain (e.g., mild headache) during the last week? [yes/no]
2. [If "yes" on #1] Where in your body did you feel pain? ________________
3. [If "yes" on #1] Where in your body was your pain the worst? ________________
4. In the last week, did you experience [respond with 100-point VAS scale where 0 is none at all and 100 is pain as bad as you can imagine, or prefer not to answer] :
    a. Vaginal pain related to childbirth or pregnancy?
    b. Pelvic pain (i.e., lower abdomen, hips) related to childbirth or pregnancy?
    c. Pain at cesarean section incision?
    d. Other pain related to childbirth or pregnancy? Describe: ________
    e. Nipple or breast pain during breastfeeding?
    f. Nipple or breast pain when not breastfeeding?
5. Have you had any of the following in the last week? [yes/no]
    a. Mastitis or breast infection
    b. Painful breast engorgement
    c. Blocked or clogged milk ducts
    d. Thrush
    e. Cracked, bleeding, or sore nipples
6. Please rate your pain at its worst during the last week. [respond with 100-point VAS scale where 0 is none at all and 100 is pain as bad as you can imagine, or prefer not to answer]
7. Please rate your pain at its least during the last week. [respond with 100-point VAS scale where 0 is none at all and 100 is pain as bad as you can imagine, or prefer not to answer]
8. Please rate your pain on average during the last week. [respond with 100-point VAS scale where 0 is none at all and 100 is pain as bad as you can imagine, or prefer not to answer]
9. What treatments or medications are you receiving for your pain?
   Treatment: _______________ If applicable, dose: ________ frequency: _______
10. During the last week, how much relief have pain treatments or medication provided? [respond with 100-point VAS scale where 0 is no relief and 100 is complete relief, or prefer not to answer]
11. During the last week, how has pain interfered with your: [respond with 100-point VAS scale where 0 is does not interfere and 100 is completely interferes, or prefer not to answer]
    a. General activity
    b. Caregiving activity
    c. Breastfeeding
    d. Mood
    e. Walking ability
    f. Normal work (includes both work outside the home and housework)
    g. Bonding with your baby
    h. Relations with other people
    i. Sleep
    j. Enjoyment of life

Modified Postpartum Stressor Scale

(Completed at baseline and then weekly starting the day after baseline until postpartum week 12, and then again at postpartum months 4 and 5. All items will be answered using a 1 to 4 Likert-type scale ranging from “Not at all stressful” to “Very stressful,” with a “not applicable” for the relationship items and a “prefer not to answer” option for all items.)

Directions: Please rate how stressful each of the following has been for you [postpartum only]: since you had your baby:

1. Relationship to spouse/partner
2. [presented to those who selected option d, e, f #5 of sociodemographics] Relationship to baby’s dad
3. Relationship to others around me
4. [postpartum only] Breastfeeding
5. [postpartum only] Being a mother
6. [postpartum only] Fussy baby
7. Financial worries
8. Work problems
9. Concerns with own health
10. Concerns about physical appearance (weight, shape)
11. Lack of sleep

Modified Epworth Sleepiness Scale

(Completed at baseline and then weekly starting the day after baseline until postpartum week 12 and then again at months 4 and 5. All items will be answered using the following response options: would never nod off, slight chance of nodding off, moderate chance of nodding off, high chance of nodding off, and a "prefer not to answer" response option.)

Directions: How likely are you to nod off or fall asleep in the following situations, in contrast to feeling just tired? This refers to how you have felt over the past week. Even if you haven't done some of these things recently, try to work out how they would have affected you. It is important that you answer each question as best you can.

1. Sitting and reading
2. Watching TV
3. Sitting inactive in a public place (e.g., in a meeting, theater, or dinner event)
4. As a passenger in a car for an hour or more without stopping for a break.
5. Lying down to rest when circumstances permit
6. Sitting and talking to someone
7. Sitting quietly after a meal without alcohol
8. In a car, while stopped for a few minutes in traffic or at a light
9. While feeding my baby
10. While rocking my baby
11. While sitting quietly with my baby

Modified Pittsburgh Sleep Quality Index
(Completed baseline, at postpartum weeks 4, 8, and 12, and then again at postpartum months 4 and 5. All items will include a "prefer not to answer" response option.)
Directions: The following questions relate to your usual sleep habits during the past week only. Your answers should indicate the most accurate reply for the majority of days and nights in the past week. Please answer all questions.

1. During the past week, what time have you usually gone to bed at night? ___
2. During the past week, how long (in minutes) has it usually taken you to fall asleep? ____
3. During the past week, how many times did you wake up each night? _____
4. During the past week, how much time did you spend awake each night? ____
5. During the past week, what time have you usually gotten up in the morning? ____
6. During the past week, how much time did you sleep during the day? ____
7. During the past week, how many total hours of actual sleep did you get at night?

For each of the remaining questions, select the best response. Please answer all questions.

8. During the past week, how often have you had trouble sleeping because you: (Response options: Not during the past week, once in the past week, twice during the past week, three or more times per week, prefer not to answer)
    a. Cannot get to sleep within 30 minutes
    b. Wake up in the middle of the night or early in the morning because of my baby
    c. Wake up in the middle of the night or early in the morning not because of my baby
    d. Have to get up to use the bathroom
    e. Cannot breathe comfortably
    f. Cough or snore loudly
    g. Feel too cold
    h. Feel too hot
    i. Have bad dreams
    j. Have pain
    k. Other reason(s), please describe: _________

    How often during the past week have you had trouble sleeping because of this? (Response options: Not during the past week, once in the past week, twice during the past week, three or more times per week, prefer not to answer)
9. During the past week, how would you rate your overall sleep quality? (Response options: very good, fairly good, fairly bad, very bad, prefer not to answer)
10. During the past week, how often have you used medicine to help you sleep (prescribed or "over the counter")? (Response options: Not during the past week, once in the past week, twice during the past week, three or more times per week, prefer not to answer)
11. During the past week, how often have you had trouble staying awake while driving, eating meals, or engaging in social activity? (Response options: Not during the past week, once in the past week, twice during the past week, three or more times per week, prefer not to answer)
12. During the past week, how much of a problem has it been to keep up enough enthusiasm to get things done? (Response options: No problem at all, only a very slight problem, somewhat of a problem, a very big problem, prefer not to answer)
13. Do you have a bed partner or room mate? (Response options: no bed partner or roommate, partner or roommate in other room, partner in same room but not same bed, partner in same bed)
14. If you had a roommate or bed partner, ask them how often in the past week you have done the following: (Response options: Not during the past week, once in the past week, twice during the past week, three or more times per week, prefer not to answer)
    a. Loud snoring
    b. Long pauses between breaths while asleep
    c. Leg twitching or jerking while you sleep
    d. Episodes of disorientation or confusion during sleep
    e. Other restlessness while you sleep (describe:____)

Modified Brief Infant Sleep Questionnaire
(Completed weekly starting at delivery and continuing until postpartum week 12, and then again at postpartum months 4 and 5. All items will include a “prefer not to answer” option.)
Considering the past seven days:

1. Where has your baby slept?
    a. Infant crib in their own room
    b. Infant crib in room with sibling
    c. Infant crib in my room
    d. In my bed
    e. Other (Describe: __________)
2. In what position does your child sleep most of the time?
    a. On their belly
    b. On their side
    c. On their back
3. How long does it take to put your baby to sleep in the evening?
    a. Hours_______
    b. Minutes______
4. How does your baby fall asleep? (Select all that apply)
    a. While feeding
    b. Being rocked
    c. Being held
    d. In bed alone
    e. In bed near parent
5. What time does your baby usually fall asleep for the night?
    a. Time:____
6. How much time does your child sleep at night (between 7 in the evening to 7 in the morning)?
    a. Hours____
    b. Minutes ______
7. How many times does your child wake up per night? _________
8. How much time during the night does your child spend in wakefulness (from 10 in the evening to 6 in the morning)
    a. Hours____
    b. Minutes ____
9. How much time does your child sleep during the day (between 7 in the morning to 8 in the evening?)
    a. Hours____
    b. Minutes ______
10. Do you consider your child’s sleep a problem?
    a. A very serious problem
    b. A small problem

Modified Father and Other Involvement

[Completed at postpartum weeks 4, 8, 12, and months 4 and 5. Note: All responses will include a “prefer not to answer” response.]

1. [At week 4 only] If your baby required specialized hospital care or admission to the NICU, how long did they stay in the hospital after birth? [weeks/days/still in hospital]
2. In the past 30 days, who has spent more than a few hours with your baby? Select all that apply.
    a. Baby’s father
    b. My partner (Note: only display if applicable)
    c. My mother
    d. A nanny or childcare provider
    e. Other (describe: ____)
3. In the past 30 days, how often has [each person selected from above] spent one or more hours a day with your baby?
    a. Every day or almost every day
    b. A few times a week
    c. A few times a month
    d. Once or twice
    e. Never
    f. I don’t know
4. In the past month, how often has [each person selected from #1 above] done the following activities with your baby? [Response options include more than once a day, once a day, a few times a week, a few times a month, rarely, not at all]
    a. Played games like “peek-a-boo” or “gotcha” with your baby
    b. Sang songs with your baby
    c. Read or looked at books with your baby
    d. Told stories to your baby
    e. Played with games or toys with your baby
    f. Got your baby get dressed
    g. Changed your baby’s diaper
    h. Gave your baby a bottle
